# COVID-19 Outbreak Prediction with Machine Learning

**DOI:** 10.1101/2020.04.17.20070094

**Authors:** Sina F. Ardabili, Amir Mosavi, Pedram Ghamisi, Filip Ferdinand, Annamaria R. Varkonyi-Koczy, Uwe Reuter, Timon Rabczuk, Peter M. Atkinson

## Abstract

Several outbreak prediction models for COVID-19 are being used by officials around the world to make informed-decisions and enforce relevant control measures. Among the standard models for COVID-19 global pandemic prediction, simple epidemiological and statistical models have received more attention by authorities, and they are popular in the media. Due to a high level of uncertainty and lack of essential data, standard models have shown low accuracy for long-term prediction. Although the literature includes several attempts to address this issue, the essential generalization and robustness abilities of existing models needs to be improved. This paper presents a comparative analysis of machine learning and soft computing models to predict the COVID-19 outbreak. Among a wide range of machine learning models investigated, two models showed promising results (i.e., multi-layered perceptron, MLP, and adaptive network-based fuzzy inference system, ANFIS). Based on the results reported here, and due to the highly complex nature of the COVID-19 outbreak and variation in its behavior from nation-to-nation, this study suggests machine learning as an effective tool to model the outbreak.

## 1. Introduction

Access to accurate outbreak prediction models is essential to obtain insights into the likely spread and consequences of infectious diseases. Governments and other legislative bodies rely on insights from prediction models to suggest new policies and to assess the effectiveness of the enforced policies [1]. The novel Coronavirus disease (COVID-19) has been reported to infect more than 2 million people, with more than 132,000 confirmed deaths worldwide. The recent global COVID-19 pandemic has exhibited a nonlinear and complex nature [2]. In addition, the outbreak has differences with other recent outbreaks, which brings into question the ability of standard models to deliver accurate results [3]. Besides the numerous known and unknown variables involved in the spread, the complexity of population-wide behavior in various geopolitical areas and differences in containment strategies had dramatically increased model uncertainty [4]. Consequently, standard epidemiological models face new challenges to deliver more reliable results. To overcome this challenge, many novel models have emerged which introduce several assumptions to modeling (e.g., adding social distancing in the form of curfews, quarantines, etc.) [5-7].

To elaborate on the effectiveness of enforcing such assumptions understanding standard dynamic epidemiological (e.g., susceptible-infected-recovered, SIR) models is essential [8]. The modeling strategy is formed around the assumption of transmitting the infectious disease through contacts, considering three different classes of well-mixed populations; susceptible to infection (class *S*), infected (class *I*), and the removed population (class *R* is devoted to those who have recovered, developed immunity, been isolated or passed away). It is further assumed that the class *I* transmits the infection to class *S* where the number of probable transmissions is proportional to the total number of contacts [9-11]. The number of individuals in the class *S* progresses as a time-series, often computed using a basic differential equation as follows:

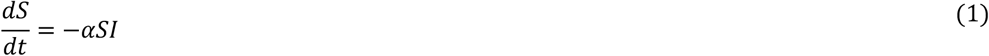

where *I* is the infected population, and *S*is the susceptible population both as fractions. *α*represents the daily reproduction rate of the differential equation, regulating the number of susceptible infectious contacts. The value of *S*in the time-series produced by the differential equation gradually declines. Initially, it is assumed that at the early stage of the outbreak *S*≈1while the number of individuals in class *I* is negligible. Thus, the increment 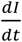 becomes linear and the class *I* eventually can be computed as follows:

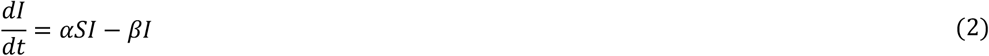

where *β*regulates the daily rate of new infections by quantifying the number of infected individuals competent in the transmission. Furthermore, the class *R*, representing individuals excluded from the spread of infection, is computed as follows:

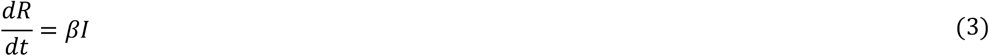

Under the unconstrained conditions of the excluded group, Eq. 3, the outbreak exponential growth can be computed as follows:

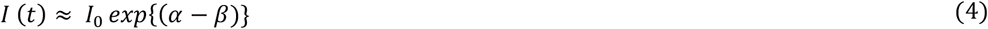

The outbreaks of a wide range of infectious diseases have been modeled using Eq. 4. However, for the COVID-19 outbreak prediction, due to the strict measures enforced by authorities, the susceptibility to infection has been manipulated dramatically. For example, in China, Italy, France, Hungary and Spain the SIR model cannot present promising results, as individuals committed voluntarily to quarantine and limited their social interaction. However, for countries where containment measures were delayed (e.g., United States) the model has shown relative accuracy [12]. Figure. 1 shows the inaccuracy of conventional models applied to the outbreak in Italy by comparing the actual number of confirmed infections and epidemiological model predictions^1^. The SEIR models through considering the significant incubation period during which individuals have been infected showed progress in improving the model accuracy for Varicella and Zika outbreak [13,14]. SEIR models assume that the incubation period is a random variable and similarly to the SIR model, there would be a disease-free-equilibrium [15,16]. It is worth mentioning that SEIR model will not work well where the parameters are non-stationary through time [17]. A key cause of non-stationarity is where the social mixing (which determines the contact network) changes through time. Social mixing determines the reproductive number *R*_0_ which is the number of susceptible individuals that an infected person will infect. Where *R*_0_ is less than 1 the epidemic will die out. Where it is greater than 1 it will spread. *R*_0_ for COVID-19 prior to lockdown was estimated as a massive 4 presenting a pandemic. It is expected that lockdown measures should bring *R*_0_ down to less than 1. the KEY reason why SEIR models are difficult to fit for COVID-19 is non-stationarity of mixing, caused by nudging (step-by-step) intervention measures.

**Figure 1.**
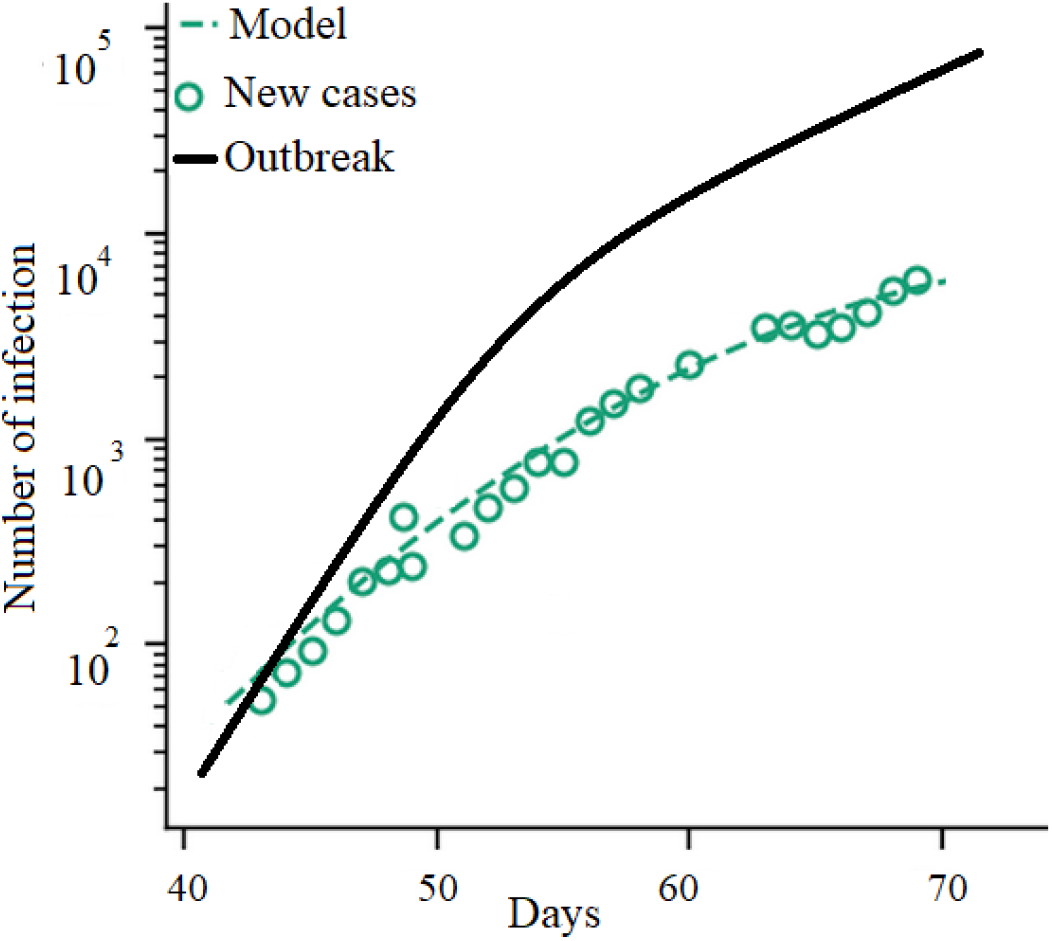
Italy’s COVID-19 outbreak: the actual number of confirmed infections vs. epidemiological model.

One can calculate that standard epidemiological models can be effective and reliable only if (a) the social interactions are stationary through time (i.e., no changes in interventions or control measures), or (b) there exists a great deal of knowledge of class *R* with which to compute Eq. 3. Often to acquire information on class *R*, several novel models included data from social media or call data records (CDR), which showed promising results [18-25]. However, observation of the behavior of COVID-19 in several countries demonstrates a high degree of uncertainty and complexity [26]. Thus, for epidemiological models to be able to deliver reliable results, they must be adapted to the local situation with an insight into susceptibility to infection [27]. This imposes a huge limit on the generalization ability and robustness of conventional models. Advancing accurate models with a great generalization ability to be scalable to model both the regional and global pandemic is, thus, essential [28].

A further drawback of conventional epidemiological models is the short lead-time. To evaluate the performance of the models, the median success of the outbreak prediction presents useful information. The median prediction factor can be calculated as follows:

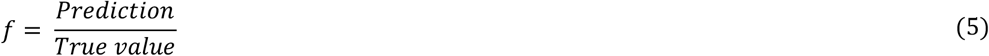

As the lead-time increases, the accuracy of the model declines. For instance, for the COVID-19 outbreak in Italy, the accuracy of the model for more than 5-days-in-the-future reduces from *f*=1for the first five days to *f*=0.86 for day 6 [12].

Due to the complexity and the large-scale nature of the problem in developing epidemiological models, machine learning (ML) has recently gained attention for building outbreak prediction models. ML approaches aim at developing models with higher generalization ability and greater prediction reliability for longer lead-times [29-33].

Although ML methods were used in modeling former pandemics (e.g., Ebola, Cholera, swine fever, H1N1 influenza, dengue fever, Zika, oyster norovirus [8,34-43]), there is a gap in the literature for peer-reviewed papers dedicated to COVID-19. Table 1 represents notable ML methods used for outbreak prediction. These ML methods are limited to the basic methods of random forest, neural networks, Bayesian networks, Naïve Bayes, genetic programming and classification and regression tree (CART). Although ML has long been established as a standard tool for modeling natural disasters and weather forecasting [44,45], its application in modeling outbreak is still in the early stages. More sophisticated ML methods (e.g., hybrids, ensembles) are yet to be explored. Consequently, the contribution of this paper is to explore the application of ML for modeling the COVID-19 pandemic. This paper aims to investigate the generalization ability of the proposed ML models and the accuracy of the proposed models for different lead-times.

**Table 1.** Notable ML methods for outbreak prediction

| Authors | Journal | Outbreak infection | Machine learning |
| --- | --- | --- | --- |
| [39] | Transboundary and Emerging Diseases | Swine fever | Random Forest |
| [35] | Geospatial Health | Dengue fever | Neural Network |
| [42] | BMC Research Notes | Influenza | Random Forest |
| [41] | Journal of Public Health Medicine | Dengue/Aedes | Bayesian Network |
| [38] | Informatica | Dengue | LogitBoost |
| [8] | Global Ecology and Biogeography | H1N1 flu | Neural Network |
| [34] | Current Science | Dengue | Adopted multi-regression and Naïve Bayes |
| [36] | Environment International | Oyster norovirus | Neural Network |
| [37] | Water Research | Oyster norovirus | Genetic programming |
| [43] | Infectious Disease Modelling | Dengue | Classification and regression tree (CART) |

The rest of this paper is organized as follows. Section two describes the methods and materials. The results are given in section three. Sections four and five present the discussion and the conclusions, respectively.

## 2. Materials and Methods

Data were collected from https://www.worldometers.info/coronavirus/country for five countries, including Italy, Germany, Iran, USA, and China on total cases over 30 days. Figure 2 presents the total case number (cumulative statistic) for the considered countries. Currently, to contain the outbreak, the governments have implemented various measures to reduce transmission through inhibiting people’s movements and social activities. Although for advancing the epidemiological models information on changes in social distancing is essential, for modeling with machine learning no assumption is required. As can be seen in Figure 2, the growth rate in China is greater than that for Italy, Iran, Germany and the USA in the early weeks of the disease.

**Figure 2.**
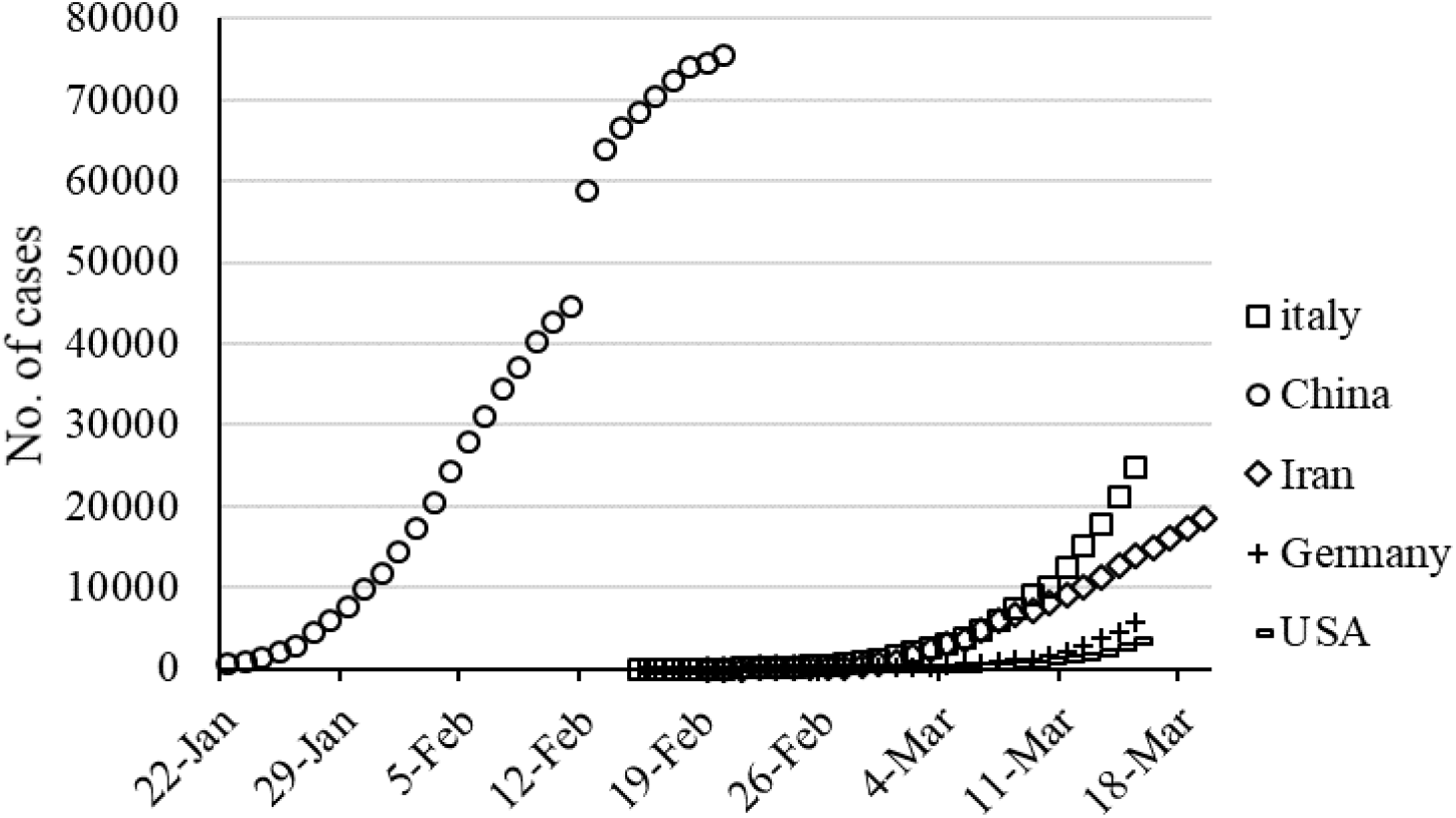
Cumulative number of cases for five countries during thirty days (https://www.worldometers.info/coronavirus/country)

The next step is to find the best model for the estimation of the time-series data. Logistic, Linear, Logarithmic, Quadratic, Cubic, Compound, Power and exponential equations (Table 2) are employed to develop the desired model.

**Table 2.** Models for mathematical forecasting

| Model description | Model name | Equation number |
| --- | --- | --- |
| $R=A/(1+\exp(((4*\mu)*(L-x)/A)+2))$ | Logistic | (6) |
| $R=Ax-B$ | Linear | (7) |
| $R=A+ B\log(x)$ | Logarithmic | (8) |
| $R=A+Bx+Cx^2$ | Quadratic | (9) |
| $R=A+Bx+Cx^2+Dx^3$ | Cubic | (10) |
| $R=AB^x$ | Compound | (11) |
| $R=Ax^B$ | Power | (12) |
| $R=AEXP(Bx)$ | Exponential | (13) |

A, B, C, µ, and L are parameters (constants) that characterize the above-mentioned functions. These constants need to be estimated to develop an accurate estimation model. One of the goals of this study was to model time-series data based on the logistic microbial growth model. For this purpose, the modified equation of logistic regression was used to estimate and predict the prevalence (i.e., *I*/Population at a given time point) of disease as a function of time. Estimation of the parameters was performed using evolutionary algorithms like GA, particle swarm optimizer, and the grey wolf optimizer. These algorithms are discussed in the following.

### Evolutionary algorithms

Evolutionary algorithms (EA) are powerful tools for solving optimization problems through intelligent methods. These algorithms are often inspired by natural processes to search for all possible answers as an optimization problem [46-48]. In the present study, the frequently used algorithms, (i.e., genetic algorithm (GA), particle swarm optimizer (PSO) and grey wolf optimizer (GWO)) are employed to estimate the parameters by solving a cost function.

#### Genetic Algorithm (GA)

GAs are considered a subset of “computational models” inspired by the concept of evolution [49]. These algorithms use “Potential Solutions” or “Candidate Solutions” or “Possible Hypotheses” for a specific problem in a “chromosome-like” data structure. GA maintains vital information stored in these chromosome data structures by applying “Recombination Operators” to chromosome-like data structures [50-53]. In many cases, GAs are employed as “Function Optimizer” algorithms, which are algorithms used to optimize “Objective Functions.” Of course, the range of applications that use the GA to solve problems is very wide [52,54]. The implementation of the GA usually begins with the production of a population of chromosomes generated randomly and bound up and down by the variables of the problem. In the next step, the generated data structures (chromosomes) are evaluated, and chromosomes that can better display the optimal solution of the problem are more likely to be used to produce new chromosomes. The degree of “goodness” of an answer is usually measured by the population of the current candidate’s answers [55-59]. The main algorithm of a GA process is demonstrated in Figure 3.

**Figure 3.**
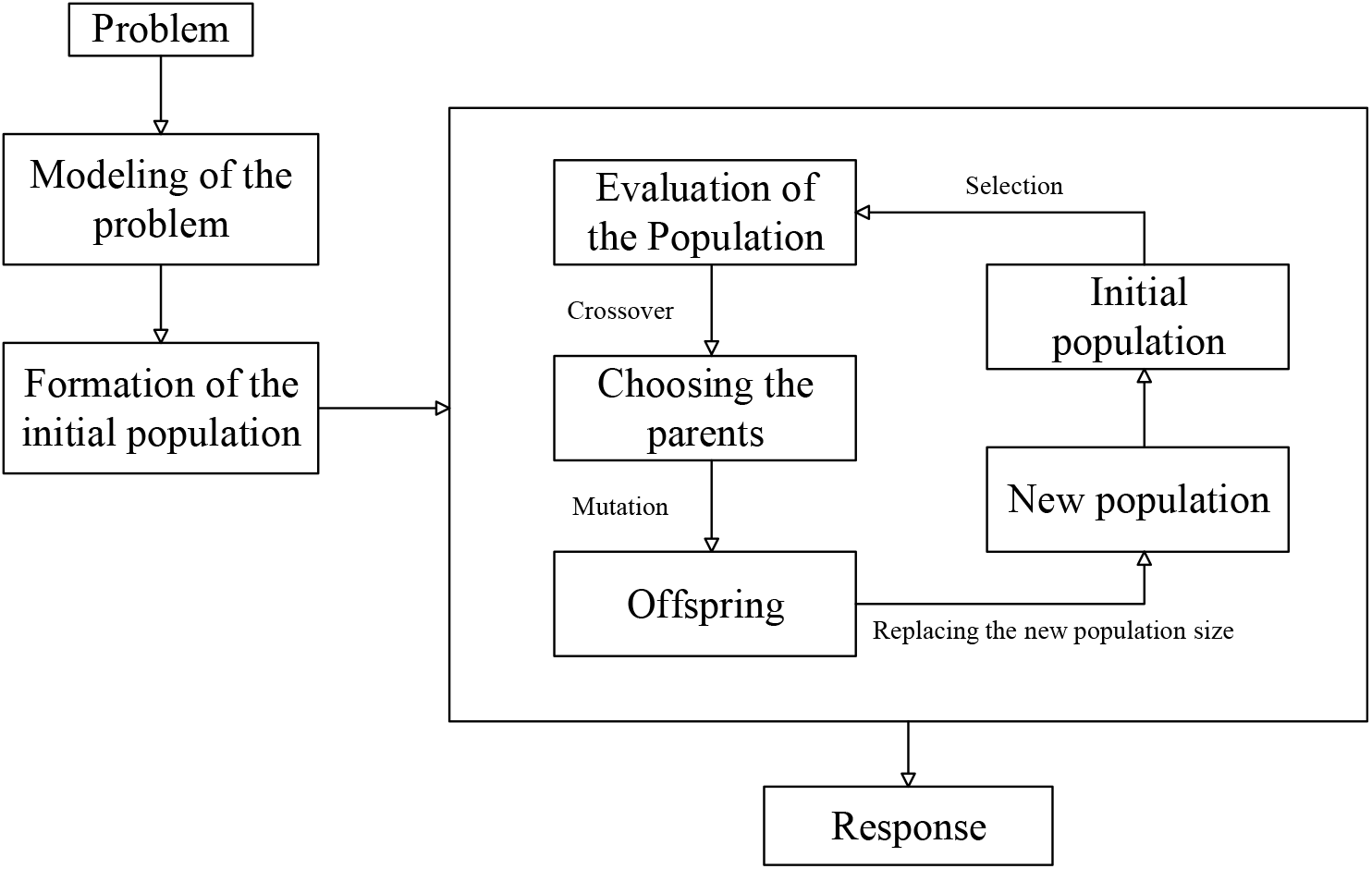
GA algorithm.

In the present study, GA [59] was employed for estimation of the parameters of Eq. 6 to 13. The population number was selected to be 300 and the maximum generation (as iteration number) was determined to be 500 according to different trial and error processes to reduce the cost function value. The cost function was defined as the mean square error between the target and estimated values according to Eq. 14:

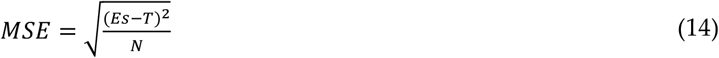

where, *Es* refer to estimated values, *T* refers to the target values and *N* refers to the number of data.

#### Particle Swarm Optimization (PSO)

In 1995, Kennedy and Eberhart [60] introduced the PSO as an uncertain search method for optimization purposes. The algorithm was inspired by the mass movement of birds looking for food. A group of birds accidentally looked for food in a space. There is only one piece of food in the search space. Each solution in PSO is called a particle, which is equivalent to a bird in the bird’s mass movement algorithm. Each particle has a value that is calculated by a competency function which increases as the particle in the search space approaches the target (food in the bird’s movement model). Each particle also has a velocity that guides the motion of the particle. Each particle continues to move in the problem space by tracking the optimal particles in the current state [60-62]. The PSO method is rooted in Reynolds’ work, which is an early simulation of the social behavior of birds. The mass of particles in nature represents collective intelligence. Consider the collective movement of fish in water or birds during migration. All members move in perfect harmony with each other, hunt together if they are to be hunted, and escape from the clutches of a predator by moving another prey if they are to be preyed upon [63-65]. Particle properties in this algorithm include [65-67]:

- Each particle independently looks for the optimal point.
- Each particle moves at the same speed at each step.
- Each particle remembers its best position in the space.
- The particles work together to inform each other of the places they are looking for.
- Each particle is in contact with its neighboring particles.
- Every particle is aware of the particles that are in the neighborhood.
- Every particle is known as one of the best particles in its neighborhood.

The PSO implementation steps can be summarized as: the first step establishes and evaluates the primary population. The second step determines the best personal memories and the best collective memories. The third step updates the speed and position. If the conditions for stopping are not met, the cycle will go to the second step.

The PSO algorithm is a population-based algorithm [68,69]. This property makes it less likely to be trapped in a local minimum. This algorithm operates according to possible rules, not definite rules. Therefore, PSO is a random optimization algorithm that can search for unspecified and complex areas. This makes PSO more flexible and durable than conventional methods. PSO deals with non-differential target functions because the PSO uses the information result (performance index or target function to guide the search in the problem area). The quality of the proposed route response does not depend on the initial population. Starting from anywhere in the search space, the algorithm ultimately converges on the optimal answer. PSO has great flexibility to control the balance between the local and overall search space. This unique PSO property overcomes the problem of improper convergence and increases the search capacity. All of these features make PSO different from the GA and other innovative algorithms [61,65,67].

In the present study, PSO was employed for estimation of the parameters of Eq. 6 to 13. The population number was selected to be 1000 and the iteration number was determined to be 500 according to different trial and error processes to reduce the cost function value. The cost function was defined as the mean square error between the target and estimated values according to Eq. 14.

#### Grey Wolf Optimizer (GWO)

One recently developed smart optimization algorithm that has attracted the attention of many researchers is the grey wolf algorithm. Like most other intelligent algorithms, GWO is inspired by nature. The main idea of the grey wolf algorithm is based on the leadership hierarchy in wolf groups and how they hunt [70]. In general, there are four categories of wolves among the herd of grey wolves, alpha, beta, delta and omega. Alpha wolves are at the top of the herd’s leadership pyramid, and the rest of the wolves take orders from the alpha group and follow them (usually there is only one wolf as an alpha wolf in each herd). Beta wolves are in the lower tier, but their superiority over Delta and omega wolves allows them to provide advice and help to alpha wolves. Beta wolves are responsible for regulating and orienting the herd based on alpha movement. Delta wolves, which are next in line for the power pyramid in the wolf herd, are usually made up of guards, elderly population, caregivers of damaged wolves, and so on. Omega wolves are also the weakest in the power hierarchy [70]. Eq. 15 to 18 are used to model the hunting tool:

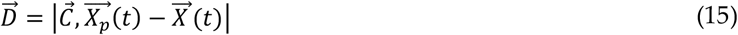

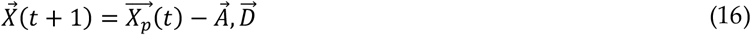

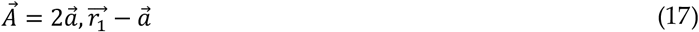

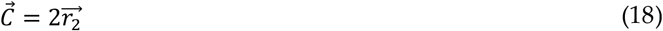

where *t* is represents repetition of the algorithm. 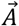 and 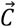 are vectors of the prey site and the 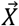 vectors represent the locations of the grey wolves. 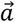 is linearly reduced from 2 to 0 during the repetition. 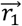 and 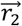 are random vectors where each element can take on realizations in the range [0.1]. The GWO algorithm flowchart is shown in Figure 4.

**Figure 4.**
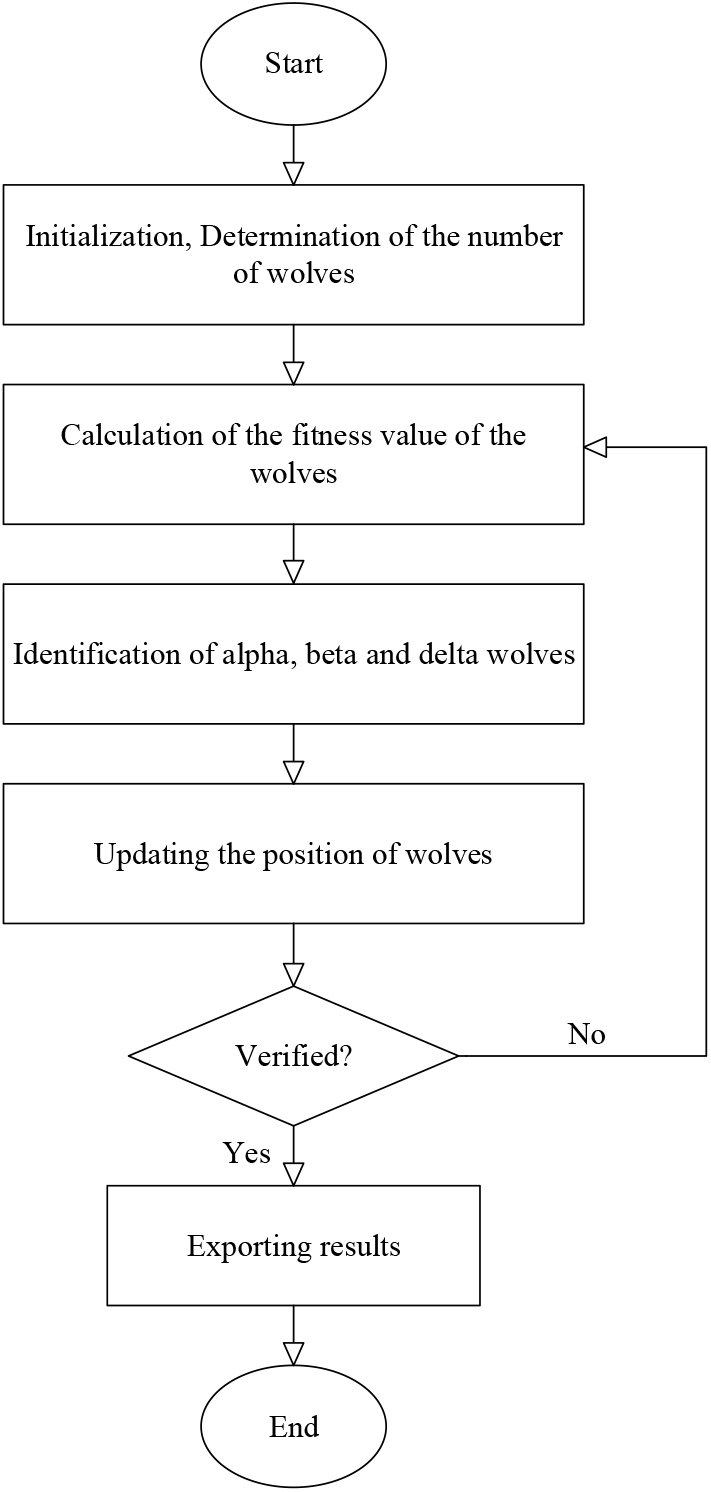
GWO algorithm.

In the present study, GWO [70] was employed for estimation of the parameters of Eq.1 to 8. The population number was selected to be 500 and the iteration number was determined to be 1000 according to different trial and error processes to reduce the cost function value. The cost function was defined as the mean square error between the target and estimated values according to Eq. 14.

### Machine learning (ML)

ML is regarded as a subset of AI. Using ML techniques, the computer learns to use patterns or “training samples” in data (processed information) to predict or make intelligent decisions without overt planning [71,72]. In other words, ML is the scientific study of algorithms and statistical models used by computer systems that use patterns and inference to perform tasks instead of using explicit instructions [73,74].

Time-series are data sequences collected over a period of time [75], which can be used as inputs to ML algorithms. This type of data reflects the changes that a phenomenon has undergone over time. Let X^t^ be a time-series vector, in which *x*_*t*_ is the outbreak at time point *t* and *T* is the set of all equidistant time points. To train ML methods effectively, we defined two scenarios, listed in Table 3.

**Table 3.** Input and output variables for training ML methods by time-series data

|  | Inputs | Input number | Output |
| --- | --- | --- | --- |
| Scenario 1 | $x_{t-1}$, $x_{t-7}$, $x_{t-14}$, and $x_{t-21}$ | Four inputs | $x_t$ (outbreak) |
| Scenario 2 | $x_{t-1}$, $x_{t-2}$, $x_{t-3}$, $x_{t-4}$, and $x_{t-5}$ | Five inputs | $x_t$ (outbreak) |

As can be seen in Table 3, scenario 1 employs data for three weeks to predict the outbreak on day *t* and scenario 2 employs outbreak data for five days to predict the outbreak for day *t*. Both of these scenarios were employed for fitting the ML methods. In the present research, two frequently used ML methods, the multi-layered perceptron (MLP) and adaptive network-based fuzzy inference system (ANFIS) are employed for the prediction of the outbreak in the five countries.

#### Multi-layered perceptron (MLP)

ANN is an idea inspired by the biological nervous system, which processes information like the brain. The key element of this idea is the new structure of the information processing system [76-78]. The system is made up of several highly interconnected processing elements called neurons that work together to solve a problem [78,79]. ANNs, like humans, learn by example. The neural network is set up during a learning process to perform specific tasks, such as identifying patterns and categorizing information. In biological systems, learning is regulated by the synaptic connections between nerves. This method is also used in neural networks [80]. By processing experimental data, ANNs transfer knowledge or a law behind the data to the network structure, which is called learning. Basically, learning ability is the most important feature of such a smart system. A learning system is more flexible and easier to plan, so it can better respond to new issues and changes in processes [81].

In ANNs, with the help of programming knowledge, a data structure is designed that can act like a neuron. This data structure is called a node [82,83]. In this structure, the network between these nodes is trained by applying an educational algorithm to it. In this memory or neural network, the nodes have two active states (on or off) and one inactive state (off or 0), and each edge (synapse or connection between nodes) has a weight. Positive weights stimulate or activate the next inactive node, and negative weights inactivate or inhibit the next connected node (if active) [78,84]. In the ANN architecture, for the neural cell *c*, the input *b*_*p*_ enters the cell from the previous cell *p. w*_*pc*_ is the weight of the input *b*_*p*_ with respect to cell *c* and *a*_*c*_ is the sum of the multiplications of the inputs and their weights [85]:

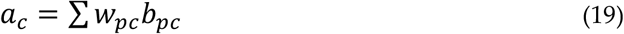

A non-linear function *Ө*_*c*_ is applied to *a*_*c*_. Accordingly, *b*_*c*_ can be calculated as Eq. 20 [85]:

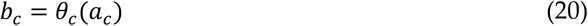

Similarly, *w*_*cn*_ is the weight of the *b*_*cn*_ which is the output of *c* to *n. W* is the collection of all the weights of the neural network in a set. For input *x* and output *y, h*_*w*_*(x)* is the output of the neural network. The main goal is to learn these weights for reducing the error values between *y* and *h*_*w*_*(x)*. That is, the goal is to minimize the cost function *Q(W)*, Eq. 21 [85]:

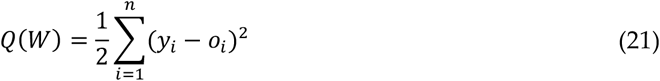

In the present research, one of the frequently used types of ANN called the MLP [76] was employed to predict the outbreak. MLP was trained using a dataset related to both scenarios (according to Table 2). For the training of the network, 8, 12, and 16 inner neurons were tried to achieve the best response. Results were evaluated by RMSE and correlation coefficient to reduce the cost function value. Figure 5 presents the architecture of the MLP.

**Figure 5.**
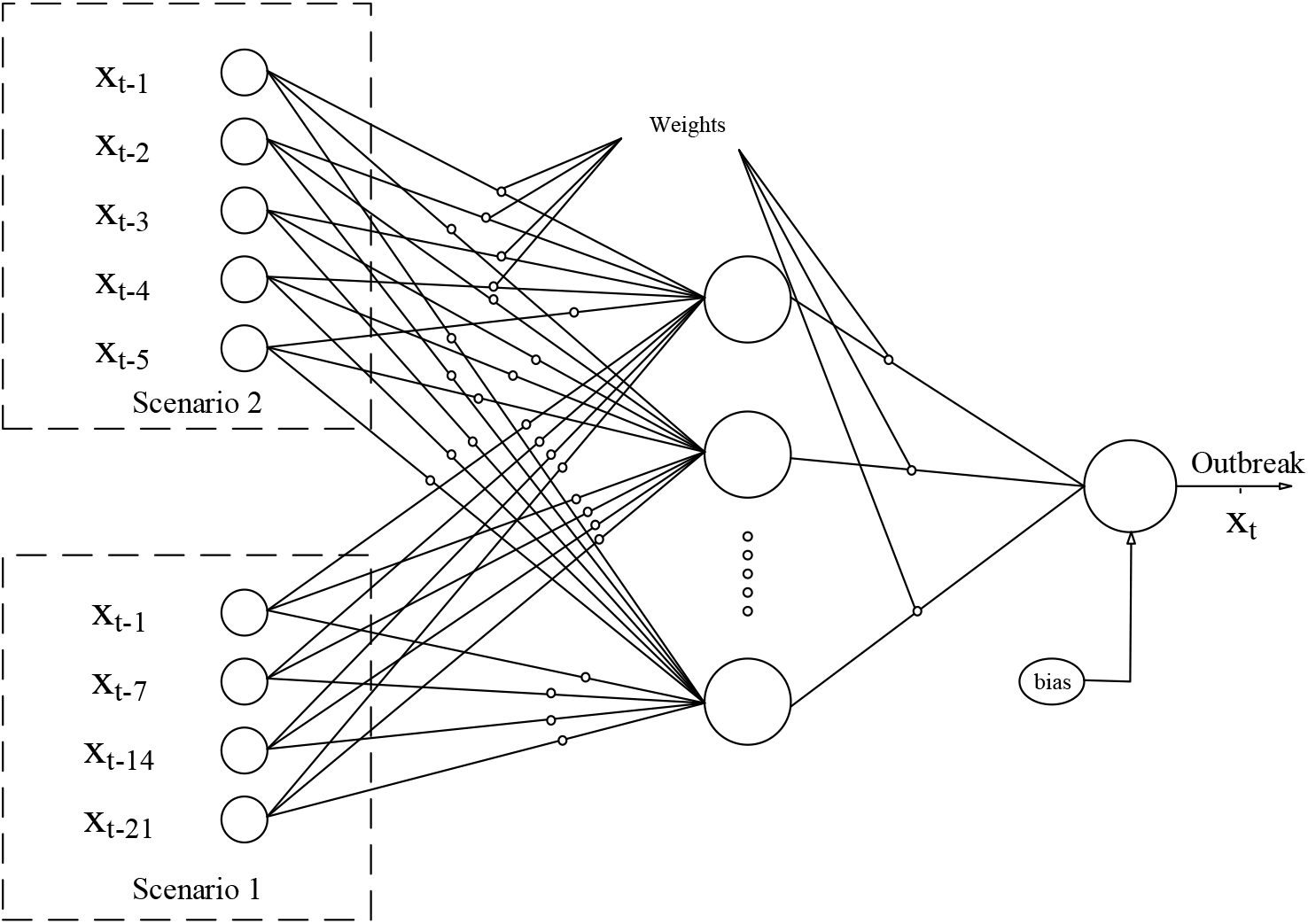
Architecture of MLP.

#### Adaptive neuro fuzzy inference system (ANFIS)

An adaptive neuro fuzzy inference system is a type of ANN based on the Takagi-Sugeno fuzzy system [86]. This approach was developed in the early 1990s. Since this system integrates the concepts of neural networks and fuzzy logic, it can take advantage of both capabilities in a unified framework. This technique is one of the most frequently used and robust hybrid ML techniques. It is consistent with a set of fuzzy if-then rules that can be learned to approximate nonlinear functions [87,88]. Hence, ANFIS was proposed as a universal estimator. An important element of fuzzy systems is the fuzzy partition of the input space [89,90]. For input *k*, the fuzzy rules in the input space make a *k* faces fuzzy cube. Achieving a flexible partition for nonlinear inversion is non-trivial. The idea of this model is to build a neural network whose outputs are a degree of the input that belongs to each class [91-93]. The membership functions (MFs) of this model can be nonlinear, multidimensional and, thus, different to conventional fuzzy systems [94-96]. In ANFIS, neural networks are used to increase the efficiency of fuzzy systems. The method used to design neural networks is to employ fuzzy systems or fuzzy-based structures. This model is a kind of division and conquest method. Instead of using one neural network for all the input and output data, several networks are created in this model:

- A fuzzy separator to cluster input-output data within multiple classes.
- A neural network for each class.
- Training neural networks with output input data in the corresponding classes. Figure 6 presents a simple architecture for ANFIS.

In the present study, ANFIS is developed to tackle two scenarios described in table 3. Each input included by two MFs with the Tri. shape; Trap. shape and Gauss. shape MFs. The output MF type was selected to be linear with a hybrid optimizer type.

**Figure 6.**
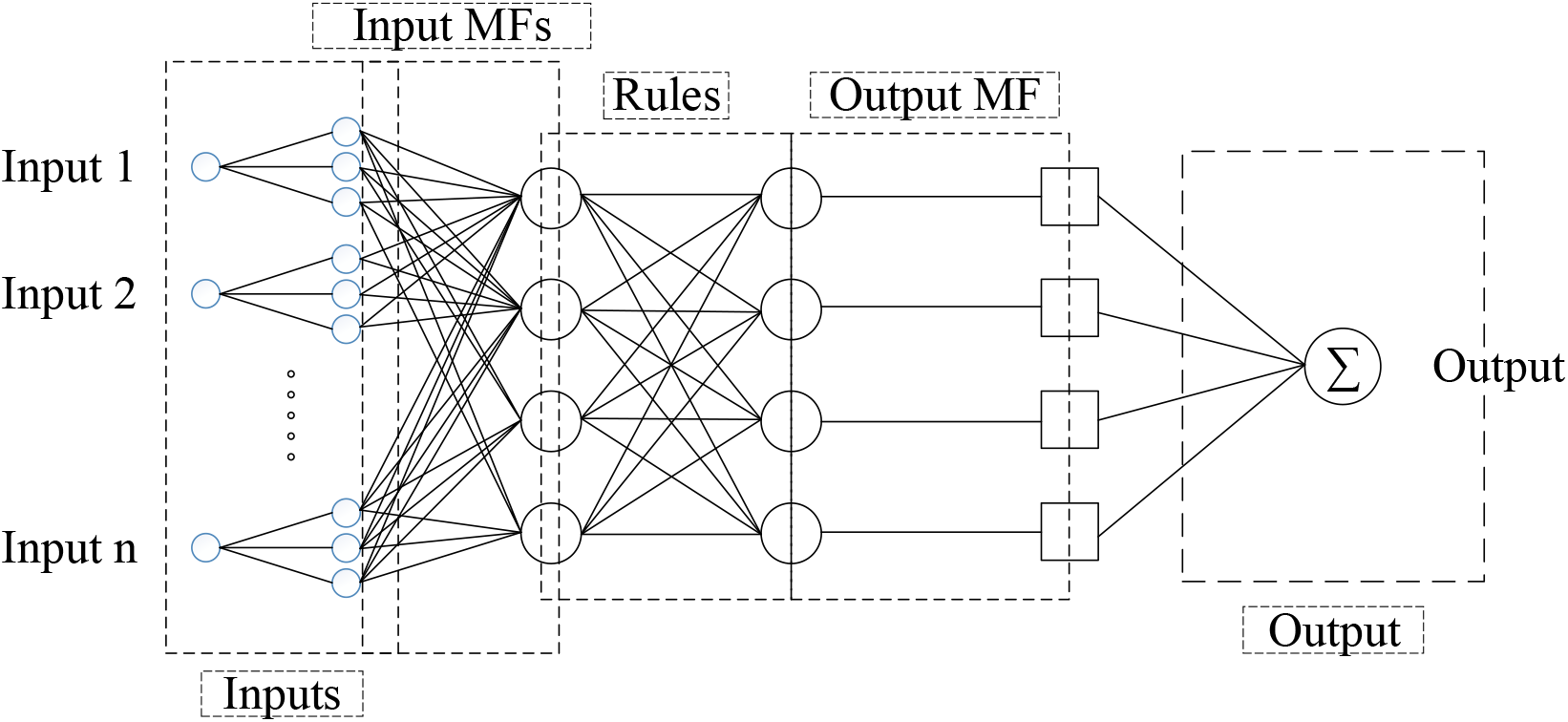
ANFIS architecture.

#### Evaluation criteria

Evaluation was conducted using the root mean square error (RMSE) and correlation coefficient. These statistics compare the target and output values and calculate a score as an index for the performance and accuracy of the developed methods [87,97]. Table 4 presents the evaluation criteria equations.

**Table 4.**
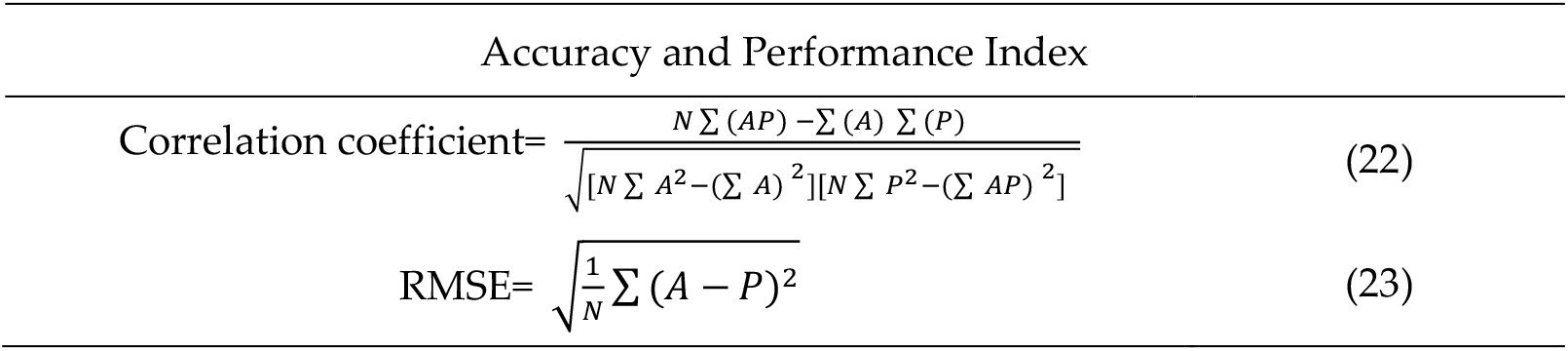
Model Evaluation metrics

Where, *N* is the number of data, *P* and *A* are, respectively, the predicted (output) and desired (target) values.

## 3. Results

Tables 5 to 12 present the results of the accuracy statistics for the logistic, linear, logarithmic, quadratic, cubic, compound, power and exponential equations, respectively. The coefficients of each equation were calculated by the three ML optimizers; GA, PSO and GWO. The table contains country name, model name, population size, number of iterations, processing time, RMSE and correlation coefficient.

**Table 5.**
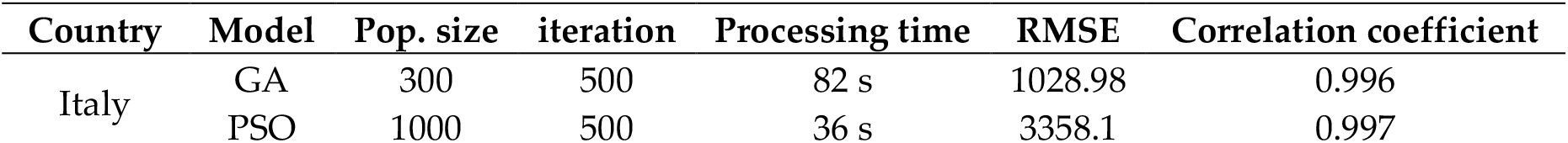
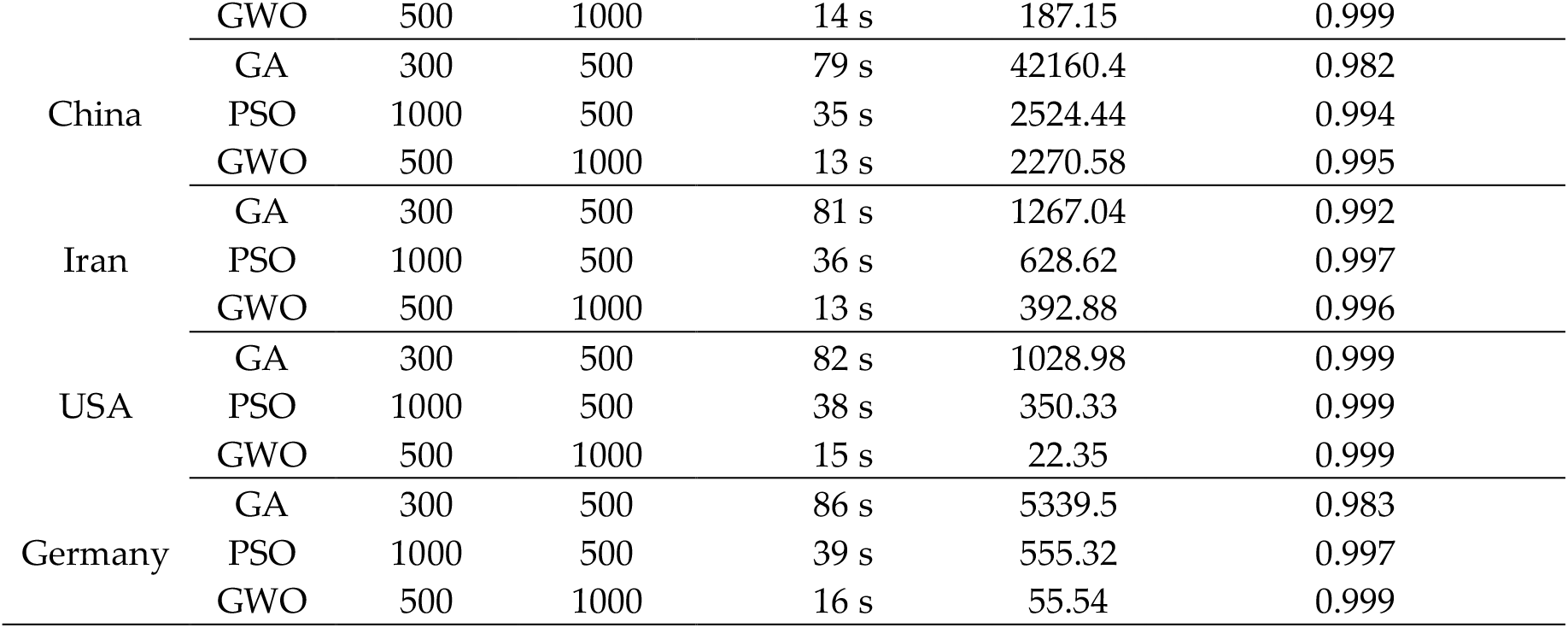
Accuracy statistics for the logistic model

**Table 6.** Accuracy statistics for the linear model

| Country | Model | Pop. size | iteration | Processing time | RMSE | Correlation coefficient |
| --- | --- | --- | --- | --- | --- | --- |
| Italy | GA | 300 | 500 | 92 s | 3774.06 | 0.845 |
|  | PSO | 1000 | 500 | 42 s | 3645.76 | 0.844 |
|  | GWO | 500 | 1000 | 16 s | 3642.44 | 0.844 |
| China | GA | 300 | 500 | 91 s | 7188.95 | 0.981 |
|  | PSO | 1000 | 500 | 39s | 6644.16 | 0.982 |
|  | GWO | 500 | 1000 | 14 s | 5039.48 | 0.982 |
| Iran | GA | 300 | 500 | 96 s | 3330.45 | 0.943 |
|  | PSO | 1000 | 500 | 45 s | 2072.71 | 0.944 |
|  | GWO | 500 | 1000 | 18 s | 1981.97 | 0.944 |
| USA | GA | 300 | 500 | 88 s | 850.22 | 0.745 |
|  | PSO | 1000 | 500 | 40 s | 596.69 | 0.746 |
|  | GWO | 500 | 1000 | 17 s | 592.48 | 0.746 |
| Germany | GA | 300 | 500 | 93 s | 1118.77 | 0.758 |
|  | PSO | 1000 | 500 | 47 s | 964.46 | 0.759 |
|  | GWO | 500 | 1000 | 20 s | 951.63 | 0.759 |

**Table 7.**
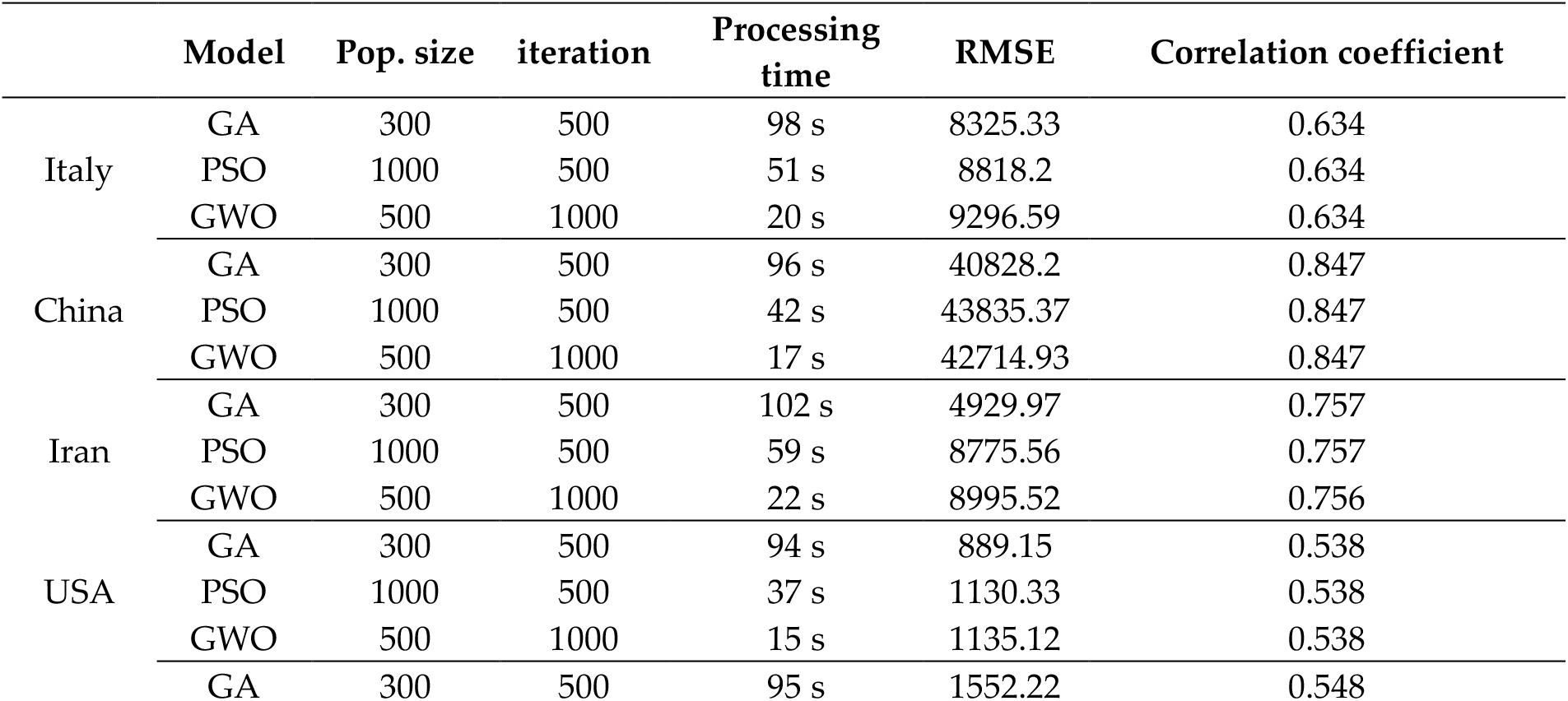
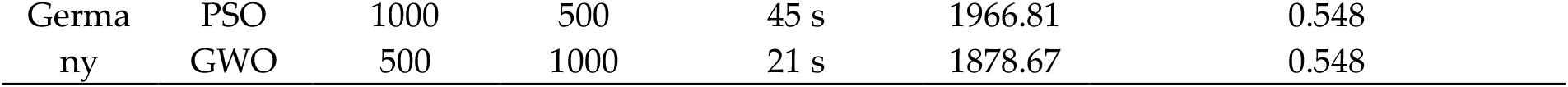
Accuracy statistics for the logarithmic model

**Table 8.** Accuracy statistics for the quadratic model

|  | Model | Pop. size | iteration | Processing time | RMSE | Correlation coefficient |
| --- | --- | --- | --- | --- | --- | --- |
| Italy | GA | 300 | 500 | 102 s | 6710.01 | 0.976 |
|  | PSO | 1000 | 500 | 54 s | 5102.4 | 0.953 |
|  | GWO | 500 | 1000 | 26 s | 1272.1 | 0.982 |
| China | GA | 300 | 500 | 100 s | 7921.33 | 0.992 |
|  | PSO | 1000 | 500 | 46 s | 4328.71 | 0.993 |
|  | GWO | 500 | 1000 | 20 s | 3710.16 | 0.993 |
| Iran | GA | 300 | 500 | 105 s | 6771.74 | 0.995 |
|  | PSO | 1000 | 500 | 62 s | 822.09 | 0.998 |
|  | GWO | 500 | 1000 | 24 s | 310.02 | 0.998 |
| USA | GA | 300 | 500 | 98 s | 754.6 | 0.931 |
|  | PSO | 1000 | 500 | 38 s | 791.92 | 0.853 |
|  | GWO | 500 | 1000 | 19 s | 307.58 | 0.938 |
| Germany | GA | 300 | 500 | 101 s | 7577 | 0.904 |
|  | PSO | 1000 | 500 | 49 s | 752.95 | 0.923 |
|  | GWO | 500 | 1000 | 26 s | 472.62 | 0.946 |

**Table 9.** Accuracy statistics for the cubic model

|  | Model | Pop. size | iteration | Processing time | RMSE | Correlation coefficient |
| --- | --- | --- | --- | --- | --- | --- |
| Italy | GA | 300 | 500 | 112 s | 7973.11 | 0.993 |
|  | PSO | 1000 | 500 | 61 s | 4827.08 | 0.996 |
|  | GWO | 500 | 1000 | 34 s | 324.33 | 0.998 |
| China | GA | 300 | 500 | 113 s | 15697.84 | 0.971 |
|  | PSO | 1000 | 500 | 59 s | 3611.15 | 0.995 |
|  | GWO | 500 | 1000 | 34 s | 2429.45 | 0.995 |
| Iran | GA | 300 | 500 | 120 s | 5852.66 | 0.995 |
|  | PSO | 1000 | 500 | 88 s | 3809.76 | 0.997 |
|  | GWO | 500 | 1000 | 39 s | 250.2 | 0.999 |
| USA | GA | 300 | 500 | 110 s | 37766.56 | 0.875 |
|  | PSO | 1000 | 500 | 49 s | 678.36 | 0.979 |
|  | GWO | 500 | 1000 | 25 s | 118.24 | 0.991 |
| Germany | GA | 300 | 500 | 116 s | 1709.06 | 0.744 |
|  | PSO | 1000 | 500 | 59 s | 1812.78 | 0.967 |
|  | GWO | 500 | 1000 | 29 s | 196.8 | 0.99 |

**Table 10.** Accuracy statistics for the compound model

|  | Model | Pop. size | iteration | Processing time | RMSE | Correlation coefficient |
| --- | --- | --- | --- | --- | --- | --- |
| Italy | GA | 300 | 500 | 92 s | 8347.51 | 0.912 |
|  | PSO | 1000 | 500 | 53 s | 195705.52 | 0.918 |
|  | GWO | 500 | 1000 | 22 s | 12585.79 | 0.951 |
| China | GA | 300 | 500 | 90 s | 41544.05 | 0.986 |
|  | PSO | 1000 | 500 | 48 s | 40195.9 | 0.988 |
|  | GWO | 500 | 1000 | 23 s | 24987.34 | 0.895 |
| Iran | GA | 300 | 500 | 99 s | 1487501.93 | 0.782 |
|  | PSO | 1000 | 500 | 81 s | 8216.81 | 0.986 |
|  | GWO | 500 | 1000 | 26 s | 13635.01 | 0.864 |
| USA | GA | 300 | 500 | 96 s | 655.62 | 0.994 |
|  | PSO | 1000 | 500 | 32 s | 1026.03 | 0.827 |
|  | GWO | 500 | 1000 | 16 s | 364.87 | 0.988 |
| Germany | GA | 300 | 500 | 98 s | 15333537.7 | 0.93 |
|  | PSO | 1000 | 500 | 72 s | 1557.23 | 0.976 |
|  | GWO | 500 | 1000 | 20 s | 431.97 | 0.998 |

**Table 11.** Accuracy statistics for the power model

|  | Model | Pop.<br>size | iteration | Processing<br>time | RMSE | Correlation<br>coefficient |
| --- | --- | --- | --- | --- | --- | --- |
| Italy | GA | 300 | 500 | 72 s | 7063.4 | 0.983 |
|  | PSO | 1000 | 500 | 40 s | 6150.52 | 0.982 |
|  | GWO | 500 | 1000 | 13 s | 3450.96 | 0.991 |
| China | GA | 300 | 500 | 65 s | 39669.92 | 0.976 |
|  | PSO | 1000 | 500 | 39 s | 19365.58 | 0.987 |
|  | GWO | 500 | 1000 | 12 s | 4078.99 | 0.989 |
| Iran | GA | 300 | 500 | 83 s | 2343032.5 | 0.951 |
|  | PSO | 1000 | 500 | 65 s | 92755.53 | 0.975 |
|  | GWO | 500 | 1000 | 15 s | 1031.6 | 0.991 |
| USA | GA | 300 | 500 | 79 s | 1030.01 | 0.779 |
|  | PSO | 1000 | 500 | 24 s | 1005.27 | 0.751 |
|  | GWO | 500 | 1000 | 11 s | 790.16 | 0.837 |
| Germany | GA | 300 | 500 | 85 s | 1475.39 | 0.871 |
|  | PSO | 1000 | 500 | 69 s | 1387.94 | 0.916 |
|  | GWO | 500 | 1000 | 14 s | 1341.91 | 0.875 |

**Table 12.**
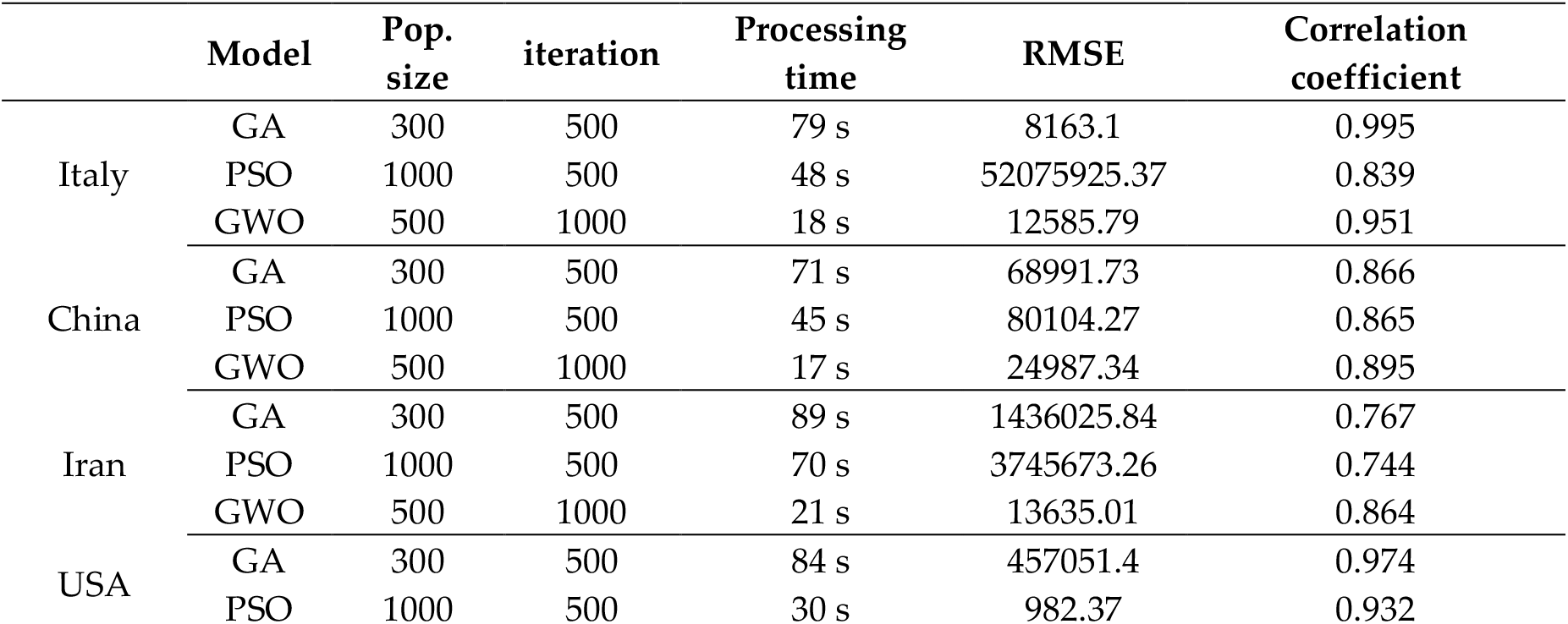
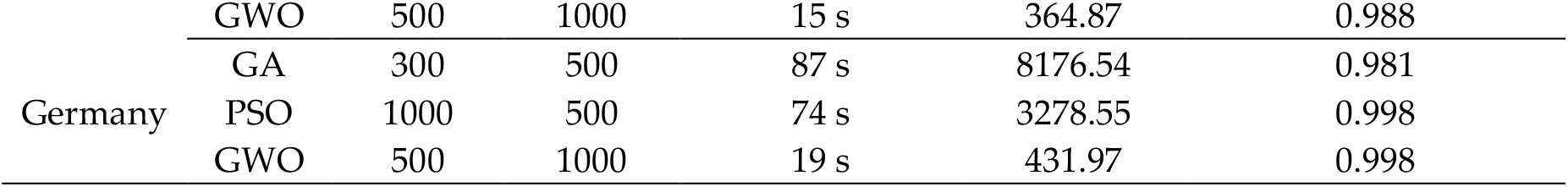
Accuracy statistics for the exponential model

According to Tables 5 to 12, GWO provided the highest accuracy (smallest RMSE and largest correlation coefficient) and smallest processing time compared to PSO and GA for fitting the logistic, linear, logarithmic, quadratic, cubic, power, compound, and exponential-based equations for all five countries. It can be suggested that GWO is a sustainable optimizer due to its acceptable processing time compared with PSO and GA. Therefore, GWO was selected as the best optimizer by providing the highest accuracy values compared with PSO and GA. In general, it can be claimed that GWO, by suggesting the best parameter values for the functions presented in Table 2, increases outbreak prediction accuracy for COVID-19 in comparison with PSO and GA. Therefore, the functions derived by GWO were selected as the best predictors for this research.

Tables 13 to 17 present the description and coefficients of the linear, logarithmic, quadratic, cubic, compound, power, exponential and logistic equations estimated by GWO. Tables 13 to 17 also present the RMSE and *r*-square values for each equation fitted to data for China, Italy, Iran, Germany and USA, respectively.

**Table 13.** Model description for China fitted by GWO **Model**

| Model name | Description | RMSE | *r*-square |
| --- | --- | --- | --- |
| Linear | $R = 3036,4 \times x - 13509,84$ | 5039.48 | 0.964 |
| Logarithmic | $R = -33948,15 + 27124,70 \times \log(x)$ | 42714.93 | 0.718 |
| Quadratic | $R = -5080,88 + 1455,98 \times x + 50,98 \times x^2$ | 3710.16 | 0.98 |
| Cubic | $R = 3984,73 - 1790,2 \times x + 308,52 \times x^2 - 5,53 \times x^3$ | 2429.45 | 0.99 |
| Compound | $R = 1601,03 \times 1.16^x$ | 24987.34 | 0.801 |
| Power | $R = 262,27 \times x^{1,69}$ | 4078.99 | 0.98 |
| Exponential | $R = 1601,03 \times \text{EXP}(0,15 \times x)$ | 24987.34 | 0.801 |
| Logistic | $R = 85011,297 / (1 + \text{EXP}(((4 \times 4483,304) * (9,423 - x) / 85011,297) + 2))$ | 2270.58 | 0.992 |

**Table 15.**
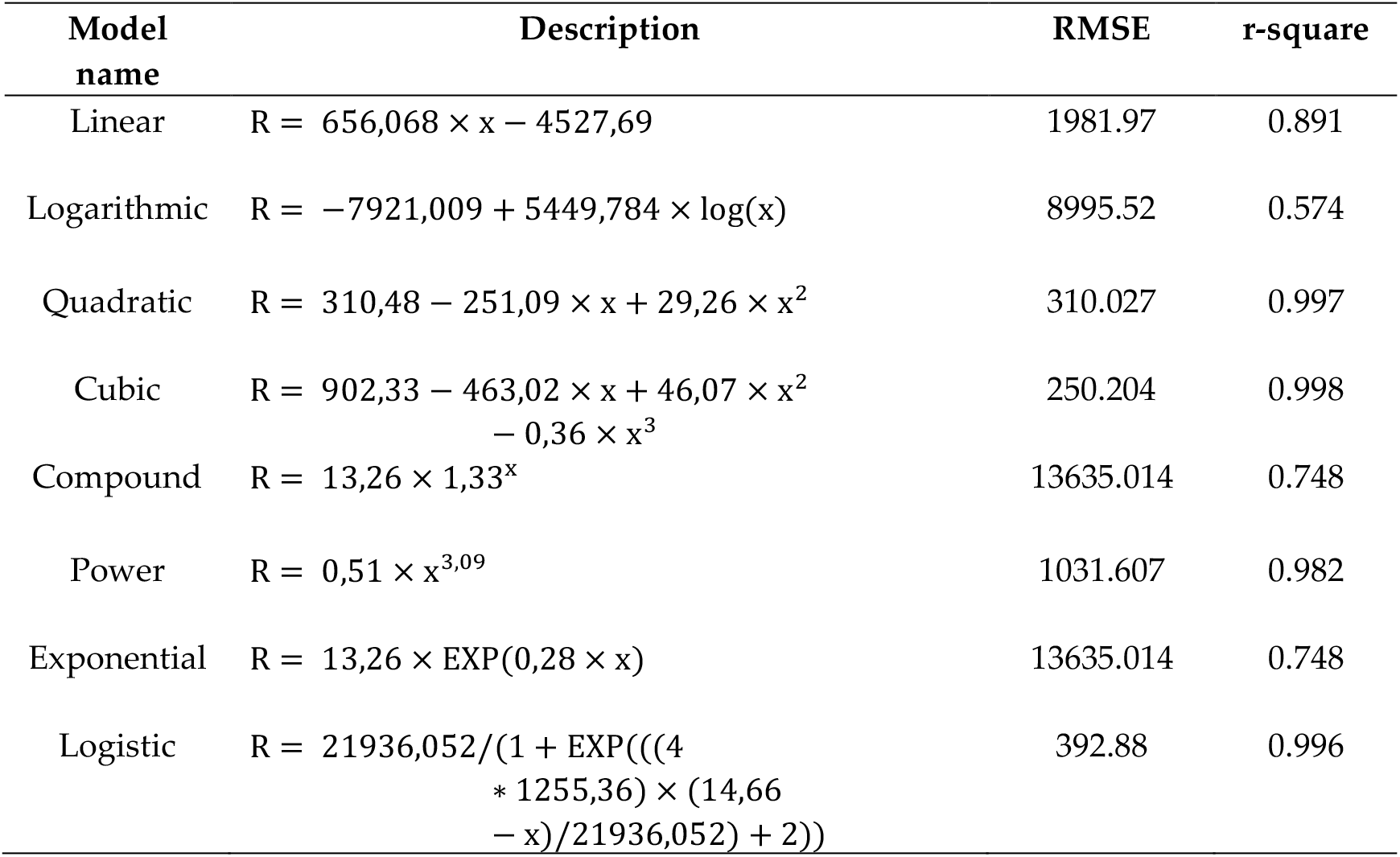
Model description for Iran fitted by GWO

**Table 16.**
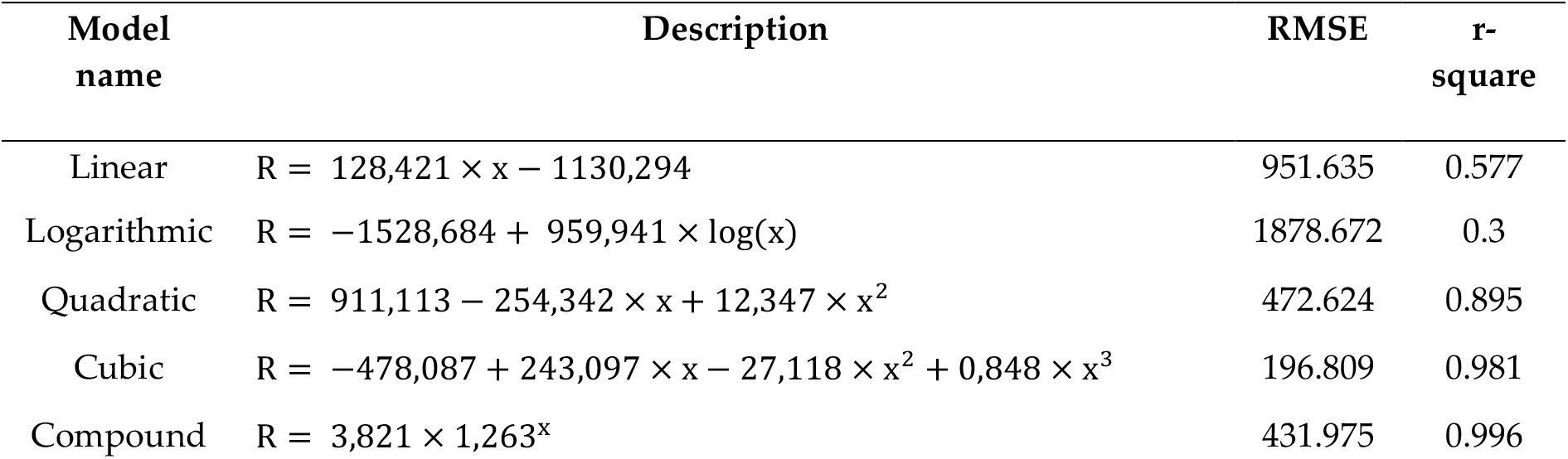
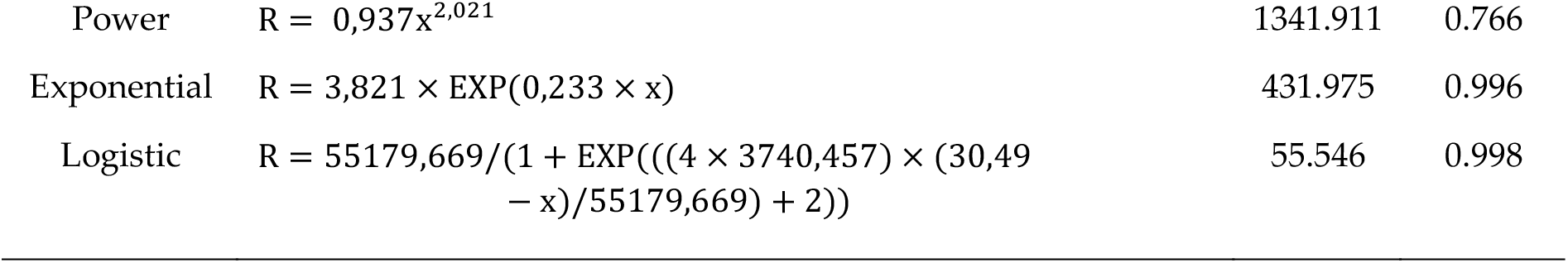
Model description for Germany fitted by GWO

**Table 17.** Model description for USA fitted by GWO

| Model name | Description | RMSE | r-square |
| --- | --- | --- | --- |
| Linear | $R = 76,833 \times x - 666,79$ | 592.486 | 0.557 |
| Logarithmic | $R = -902,637 + 573,32 \times \log(x)$ | 1135.124 | 0.289 |
| Quadratic | $R = 584,76 - 157,831 \times x + 7,569 \times x^2$ | 307.585 | 0.88 |
| Cubic | $R = -333,235 + 170,881 \times x - 18,509 \times x^2 + 0,56 \times x^3$ | 118.247 | 0.982 |
| Compound | $R = 6,296 \times 1,214^x$ | 364.875 | 0.977 |
| Power | $R = 1,707 \times x^{1,735}$ | 790.163 | 0.702 |
| Exponential | $R = 6,296 \times \text{EXP}(0,194 \times x)$ | 364.875 | 0.977 |
| Logistic | $R = 32604,552 / (1 + \text{EXP}(((4 \times 2288,932) \times (30,303 - x) / 32604,552) + 2))$ | 22.354 | 0.999 |

As is clear from Tables 13 to 17, in general, the logistic equation followed by the quadratic and cubic equations provided the smallest RMSE and the largest *r*-square values for the prediction of COVID-19 outbreak. The claim can also be considered from Figure 7 to 11, which presents the capability and trend of each model derived by GWO in the prediction of COVID-19 cases for China, Italy, Iran, Germany, and the USA, respectively.

**Figure 7.**
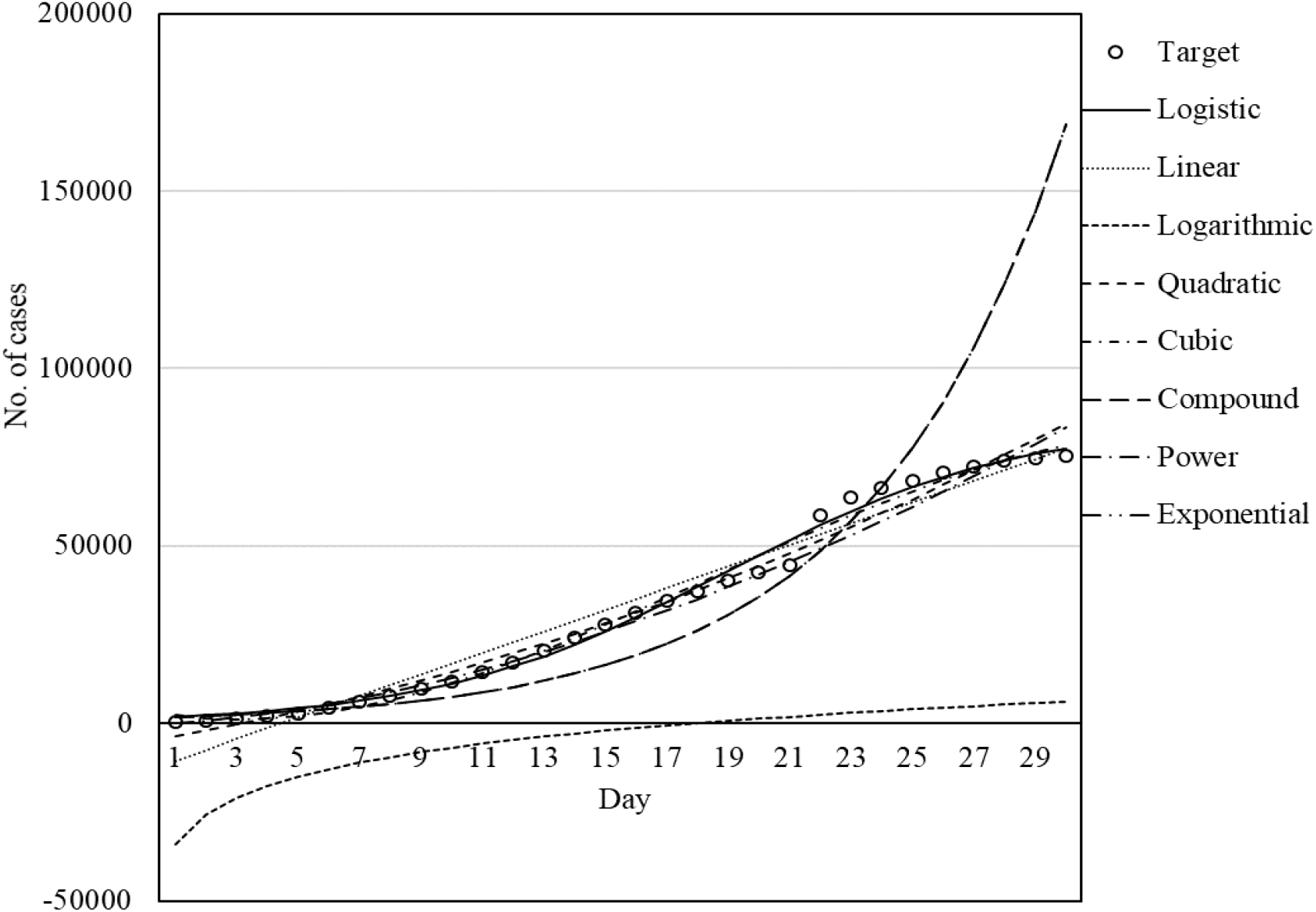
Fitness graph for China fitted by GWO.

Figures 7 to 11 illustrate the fit of the models investigated in this paper. The best fit for the prediction of COVID-19 cases was achieved for the logistic model followed by cubic and quadratic models for China (Figure 7), logistic followed by cubic models for Italy (Figure 8), cubic followed by logistic and quadratic models for Iran (Figure 9), the logistic model for Germany (Figure 10), and logistic model for the USA (Figure 11).

**Figure 8.**
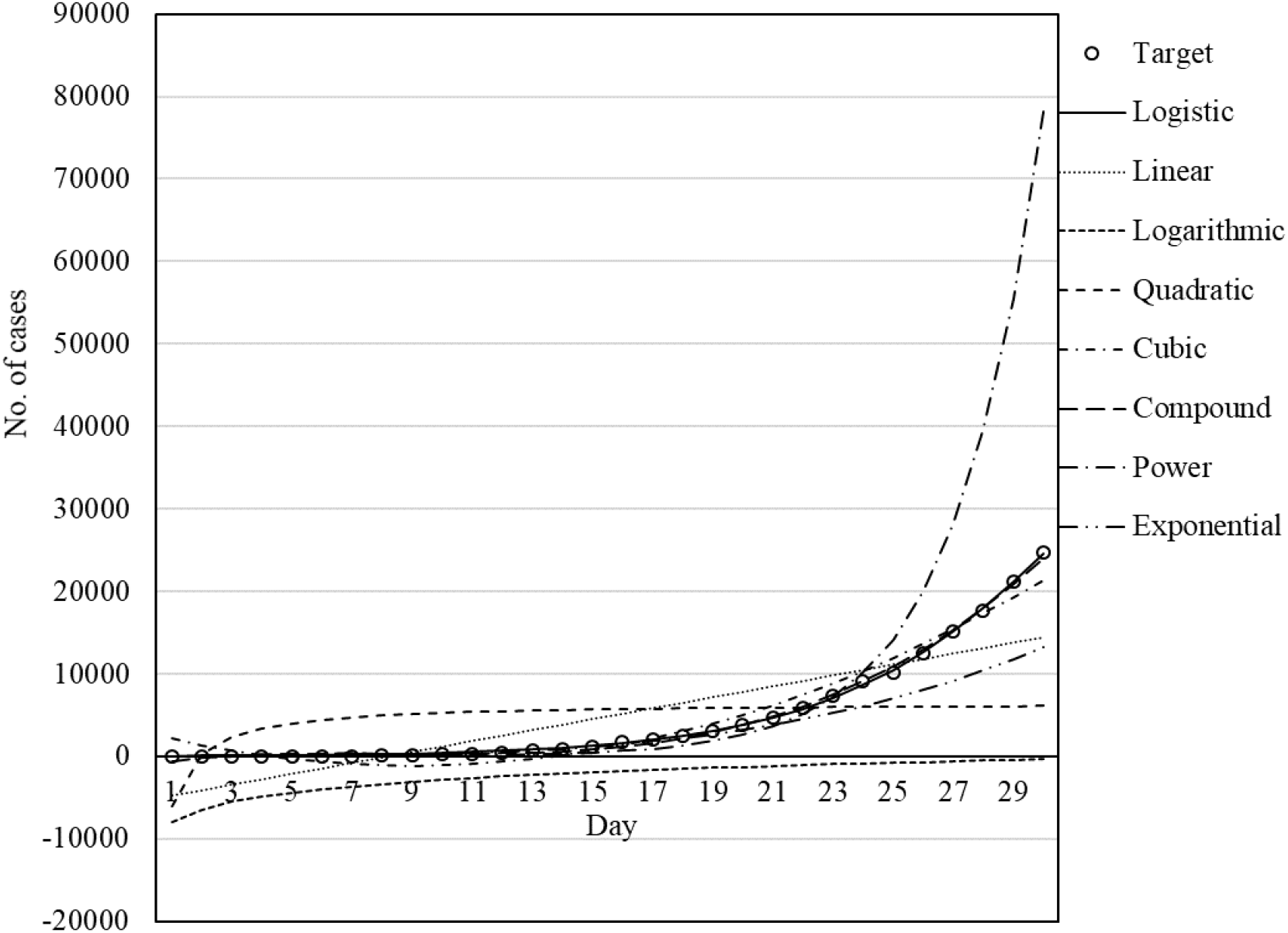
Set of models for Italy fitted by GWO.

**Figure 9.**
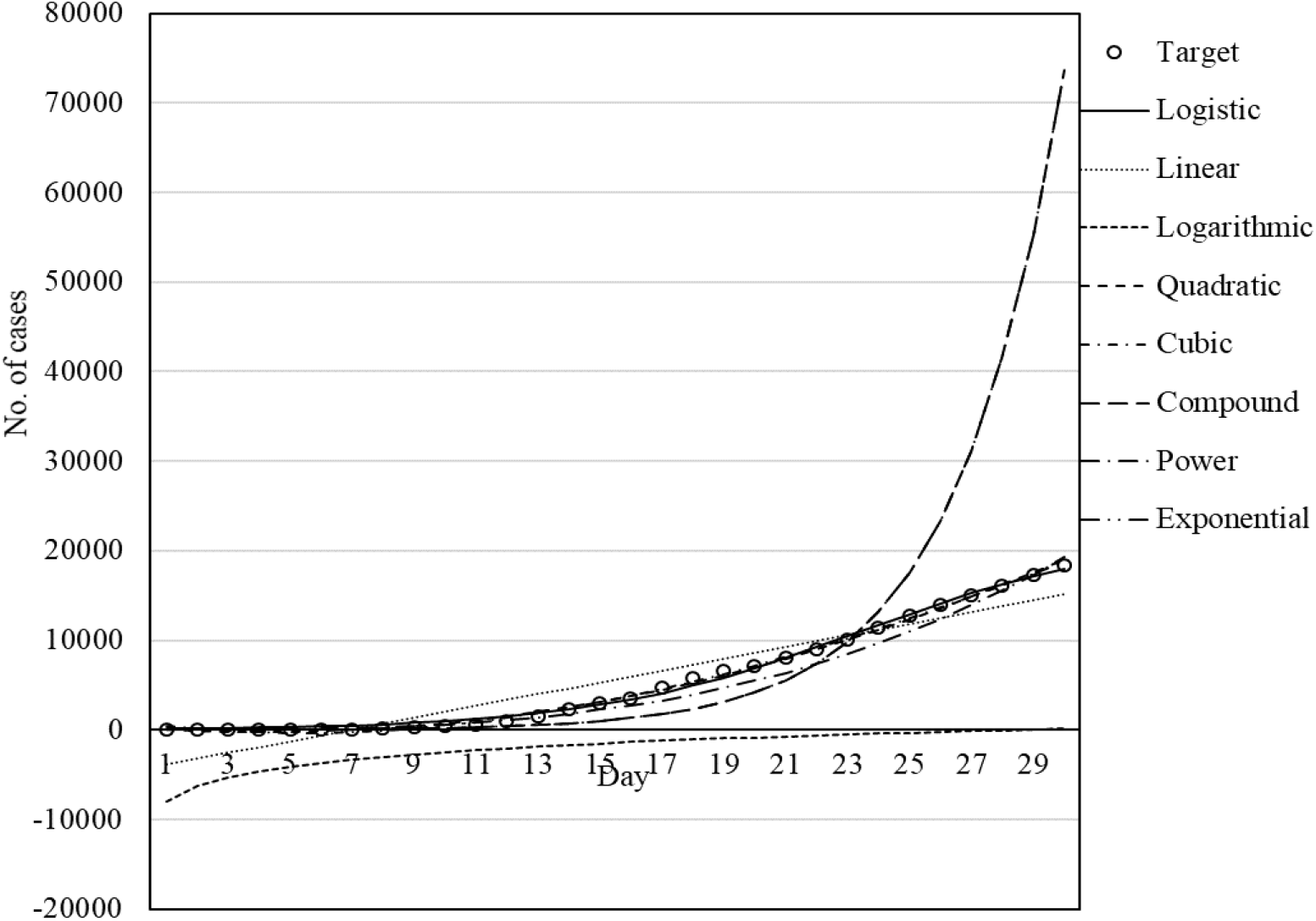
Set of models for Iran fitted by GWO.

**Figure 10.**
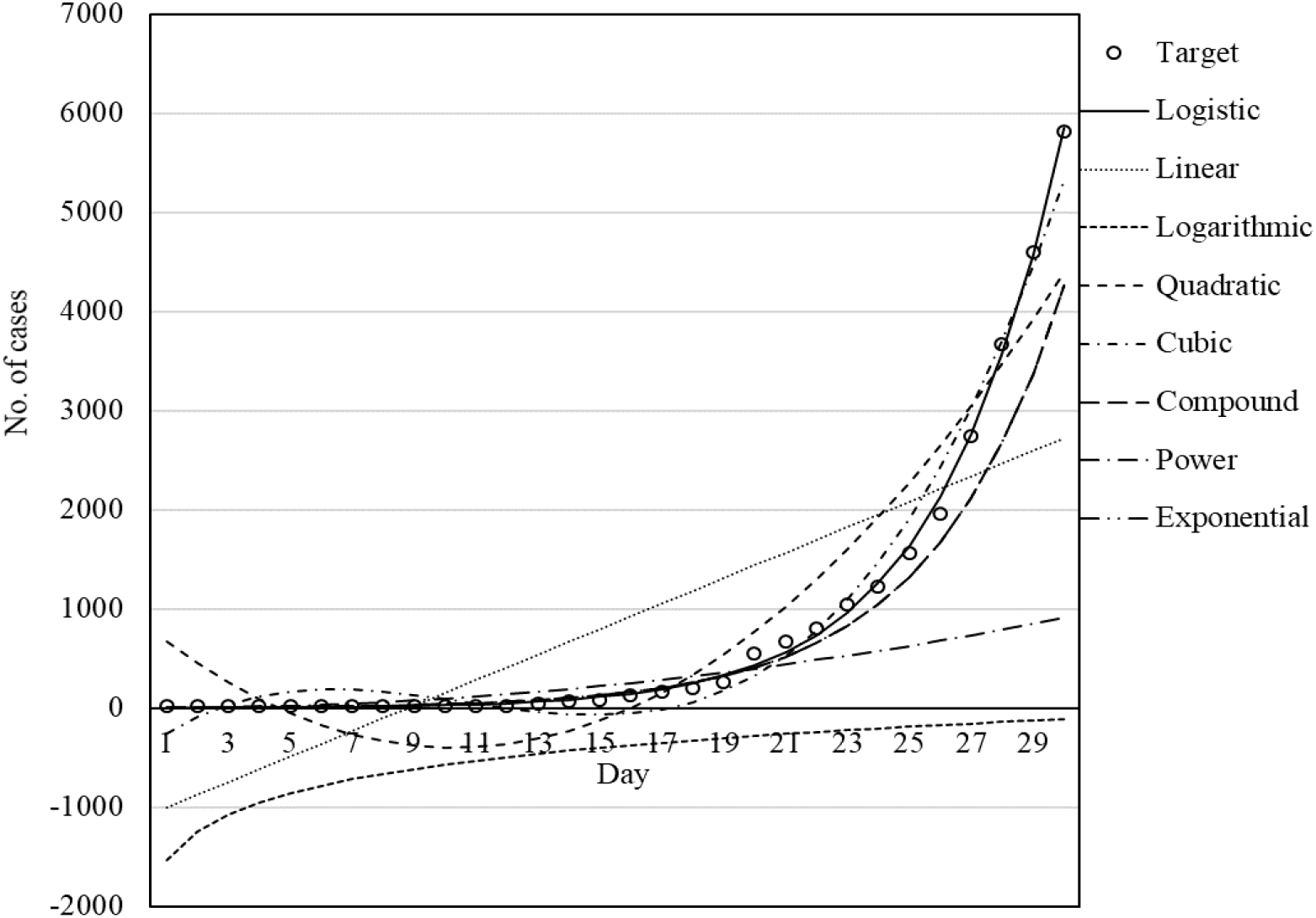
Set of models for Germany fitted by GWO.

**Figure 11.**
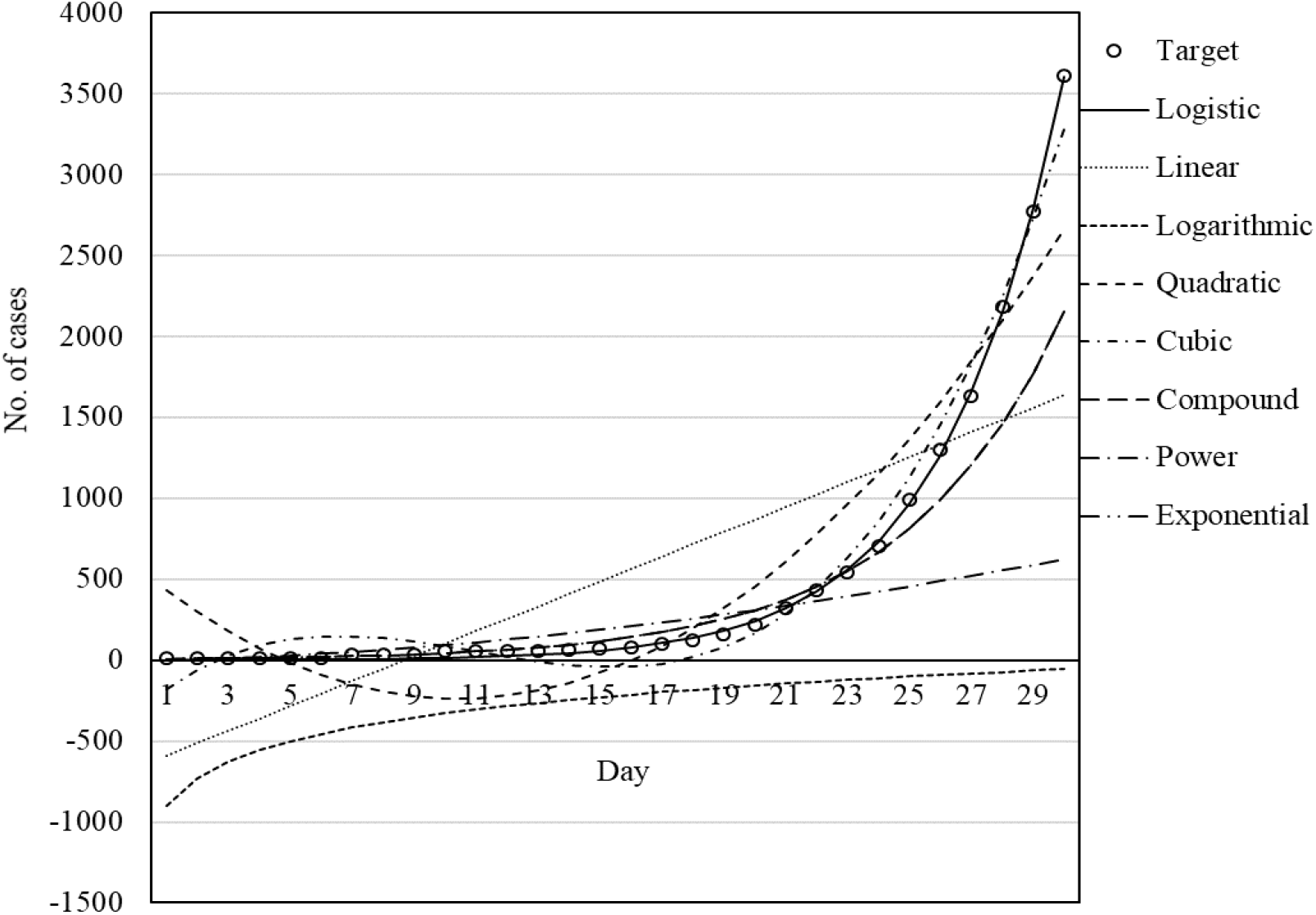
Set of models for USA fitted by GWO.

### Machine learning results

This section presents the results for the training stage of ML methods. MLP and ANFIS were employed as single and hybrid ML methods, respectively. ML methods were trained using two datasets related to scenario 1 and scenario 2. Table 14 presents the results of the training phase.

**Table 14.** Model description for Italy fitted by GWO

| Model name | Description | RMSE | *r*-square |
| --- | --- | --- | --- |
| Linear | $R = 663,71 \times x - 5437,25$ | 3642.44 | 0.713 |
| Logarithmic | $R = -7997,93 + 5162,83 \times \log(x)$ | 9296.59 | 0.402 |
| Quadratic | $R = 2998,21 - 917,93 \times x + 51,02 \times x^2$ | 1272.1 | 0.965 |
| Cubic | $R = -978,55 + 506,05 \times B2 - 61,95 \times x^2 + 2,42 \times x^3$ | 324.33 | 0.997 |
| Compound | $R = 2,78 \times 1,406^x$ | 12585.79 | 0.904 |
| Power | $R = 0,096 \times x^{3,476}$ | 3450.96 | 0.984 |
| Exponential | $R = 2,786 \times \text{EXP}(0,341 \times x)$ | 12585.79 | 0.904 |
| Logistic | $R = 70731,084 / (1 + \text{EXP}(((4 \times 3962,88) \times (23,88 - x) / 70731,08) + 2))$ | 187.15 | 0.999 |

According to Table 18, the dataset related to scenarios 1 and 2 have different performance values. Accordingly, for Italy, the MLP with 16 neurons provided the highest accuracy for scenario 1 and ANFIS with tri. MF provided the highest accuracy for scenario 2. By considering the average values of the RMSE and correlation coefficient, it can be concluded that scenario 1 is more suitable for modeling outbreak cases in Italy, as it provides a higher accuracy (the smallest RMSE and the largest correlation coefficient) than scenario 2.

**Table 18.** Results for the training phase of the ML methods

|  | Scenario 1 |  |  |  |  |  | Scenario2 |  |  |  |  |  |
| --- | --- | --- | --- | --- | --- | --- | --- | --- | --- | --- | --- | --- |
|  | MLP |  |  | ANFIS |  |  | MLP |  |  | ANFIS |  |  |
|  | No. of neurons | r | RMSE | MF type | r | RMSE | No. of neurons | r | RMSE | MF type | r | RMSE |
| Italy | 8 | 0.999 | 190.81 | Tri. | 0.999 | 189.76 | 8 | 0.999 | 199.52 | **Tri.** | **0.999** | **188.55** |
|  | 12 | 0.999 | 194.84 | Trap. | 0.841 | 3743.63 | 12 | 0.999 | 195.79 | Trap. | 0.876 | 3276 |
|  | **16** | **0.999** | **188.18** | Gauss | 0.998 | 320.93 | 16 | 0.999 | 195.2 | Gauss | 0.999 | 206.66 |
| **Average** |  | **0.999** | **191.27** |  | **0.946** | **1418.1** | **Average** | **0.999** | **196.83** |  | **0.958** | **1223.73** |
| China | 8 | 0.995 | 2287.55 | Tri. | 0.996 | 2293.09 | 8 | 0.996 | 2265.95 | Tri. | 0.996 | 2272.13 |
|  | 12 | 0.996 | 2259.95 | Trap. | 0.987 | 4231.05 | 12 | 0.996 | 2285.73 | Trap. | 0.989 | 3835.34 |
|  | 16 | 0.995 | 2407.16 | Gauss | 0.996 | 2358.3 | 16 | 0.996 | 2260.05 | Gauss | 0.996 | 2272.58 |
|  | **Average** | **0.995** | **2318.22** |  | **0.993** | **2960.81** | **Average** | **0.996** | **2270.57** |  | **0.993** | **2793.35** |
| Iran | 8 | 0.998 | 392.17 | Tri. | 0.998 | 395.33 | 8 | 0.998 | 404.21 | Tri. | 0.998 | 394.04 |
|  | 12 | 0.998 | 391.04 | Trap. | 0.977 | 1282.33 | 12 | 0.998 | 392.77 | Trap. | 0.986 | 994 |
|  | 16 | 0.998 | 392.19 | Gauss | 0.998 | 396.51 | 16 | 0.998 | 395.43 | **Gauss** | **0.998** | **391.96** |
|  | **Average** | **0.998** | **391.8** |  | **0.991** | **391.39** | **Average** | **0.998** | **397.47** |  | **0.994** | **593.33** |
| Germany | 8 | 0.999 | 55.6 | Tri. | 0.999 | 56.25 | 8 | 0.999 | 55.58 | Tri. | 0.999 | 55.63 |
|  | 12 | 0.999 | 55.38 | Trap. | 0.12 | 1658.7 | 12 | 0.999 | 55.56 | Trap. | 0.13 | 1537.26 |
|  | 16 | 0.999 | 55.58 | Gauss | 0.998 | 154.99 | 16 | 0.999 | 55.56 | Gauss | 0.999 | 62.91 |
|  | **Average** | **0.999** | **55.52** |  | **0.705** | **623.31** | **Average** | **0.999** | **55.56** |  | **0.709** | **551.93** |
| USA | 8 | 0.999 | 21.65 | Tri. | 0.999 | 21.75 | 8 | 0.999 | 22.31 | Tri. | 0.999 | 22.52 |
|  | 12 | 0.999 | 22.36 | Trap. | 0.22 | 861.08 | 12 | 0.999 | 22.3 | Trap. | 0.2 | 935.41 |
|  | 16 | 0.999 | 22.31 | Gauss | 0.998 | 86.32 | 16 | 0.999 | 22.4 | Gauss | 0.999 | 25.03 |
|  | **Average** | **0.999** | **22.1** |  | **0.739** | **323.05** | **Average** | **0.999** | **22.33** |  | **0.739** | **327.65** |

For the dataset related to China, for both scenarios, MLP with 12 and 16 neurons, respectively for scenarios 1 and 2, provided the highest accuracy compared with the ANFIS model. By considering the average values of RMSE and correlation coefficient, it can be concluded that scenario 2 with a larger average correlation coefficient and smaller average RMSE than scenario 1 is more suitable for modeling the outbreak in China.

For the dataset of Iran, MLP with 12 neurons in the hidden layer for scenario 1 and ANFIS with Gaussian MF type for scenario 2 provided the best performance for the prediction of the outbreak. By considering the average values of the RMSE and correlation coefficient, it can be concluded that scenario 1 provided better performance than scenario 2. Also, in general, the MLP has higher prediction accuracy compared with the ANFIS method.

In Germany, MLP with 12 neurons in its hidden layer provided the highest accuracy (smallest RMSE and largest correlation coefficient). By considering the average values of the RMSE and correlation coefficient, it can be concluded that scenario 1 is more suitable for the prediction of the outbreak in Germany than scenario 2.

In the USA, the MLP with 8 and 12 neurons, respectively, for scenarios 1 and 2, provided higher accuracy (the smallest RMSE and the largest correlation coefficient values) than the ANFIS model. By considering the average values of the RMSE and correlation coefficient values, it can be concluded that scenario 1 is more suitable than scenario 2, and MLP is more suitable than ANFIS for outbreak prediction.

Figures 12 to 16 present the model fits for Italy, China, Iran, Germany, and the USA, respectively. By comparing Figure 12 to 16 with Figures 7 to 11, it can be concluded that the MLP and the logistic model fitted by GWO provided a better fit than the other models. In addition, the ML methods provided better performance compared with other models.

**Figure 12.**
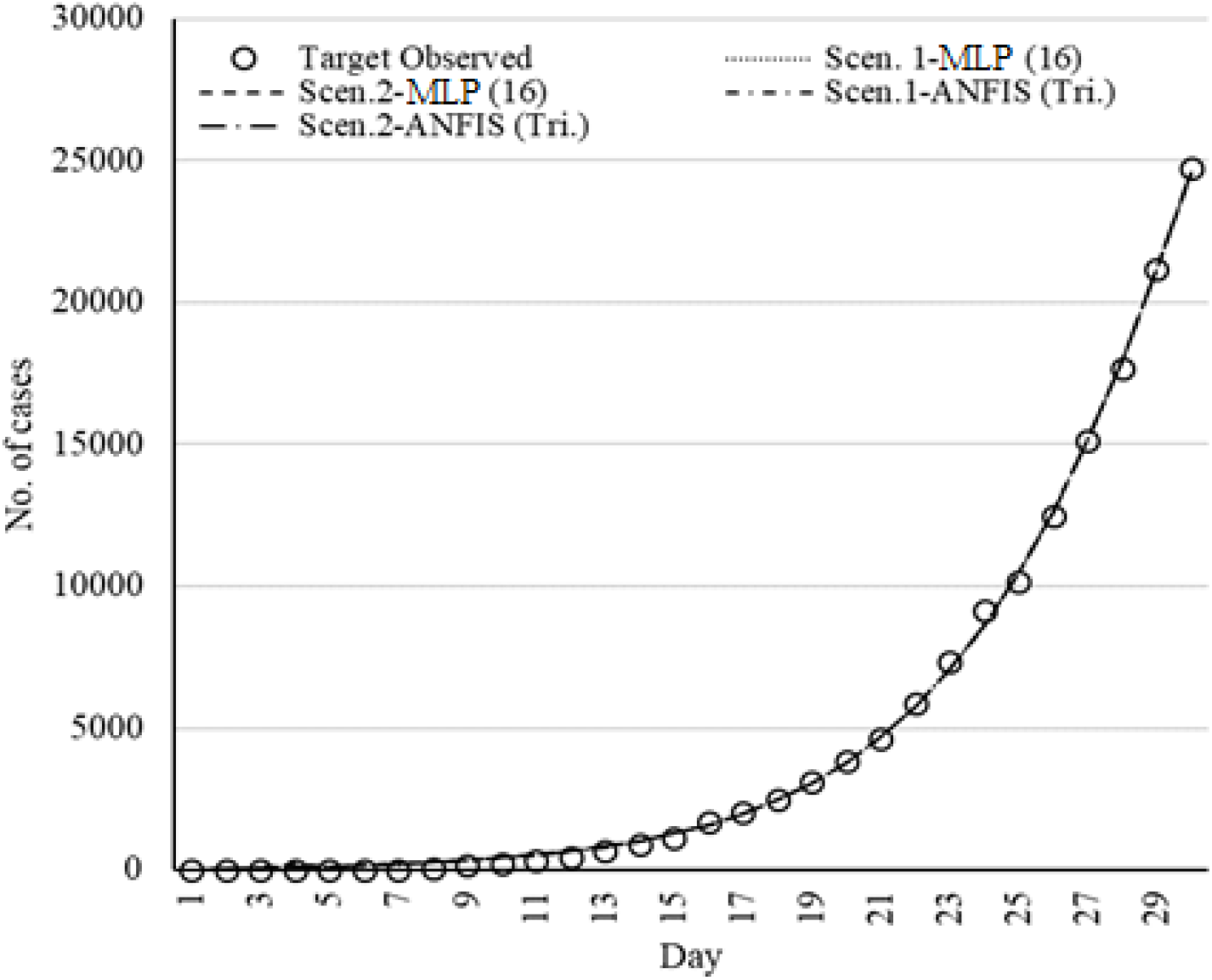
Set of models for Italy fitted by ML methods.

**Figure 13.**
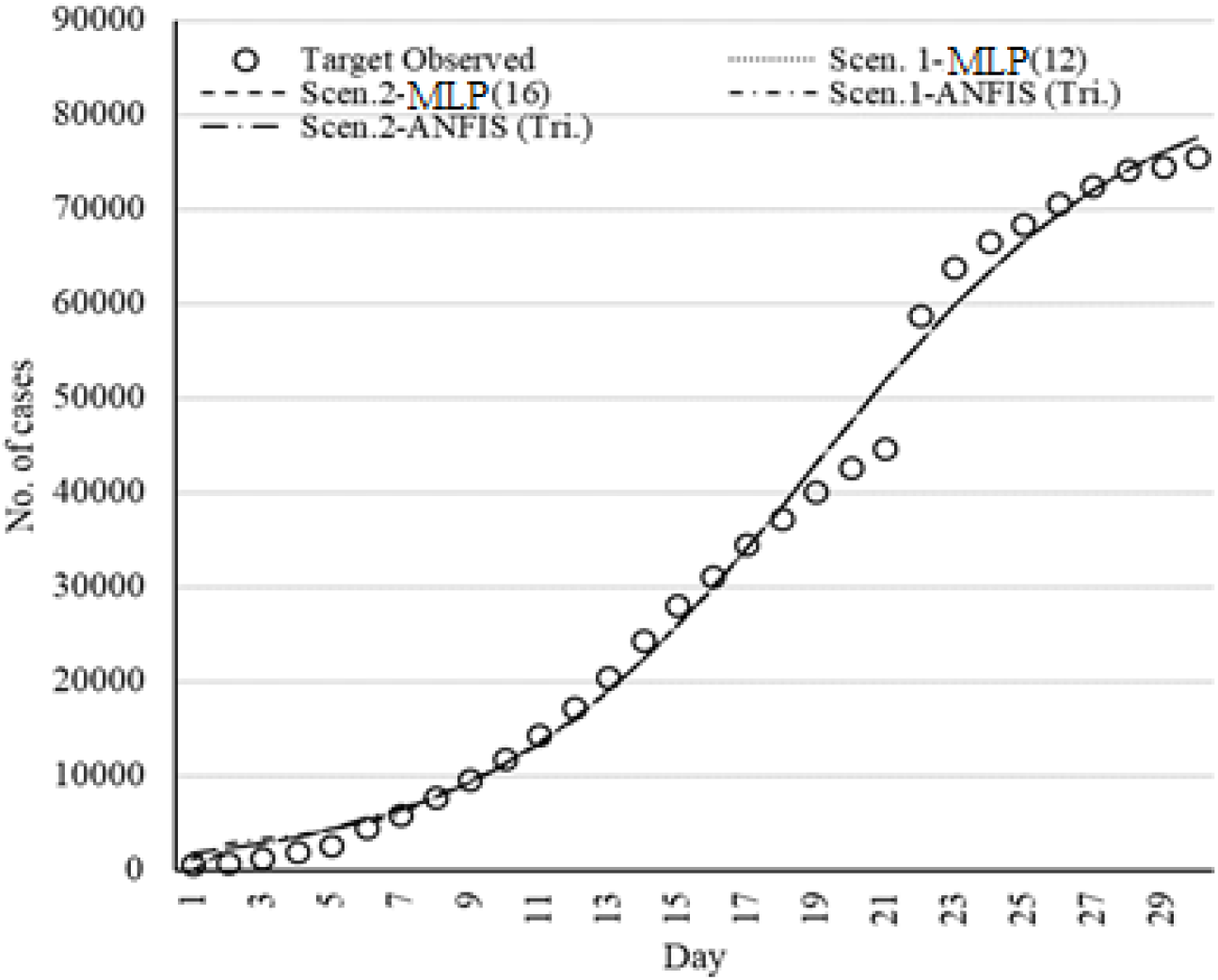
Set of models for China fitted by ML methods.

**Figure 14.**
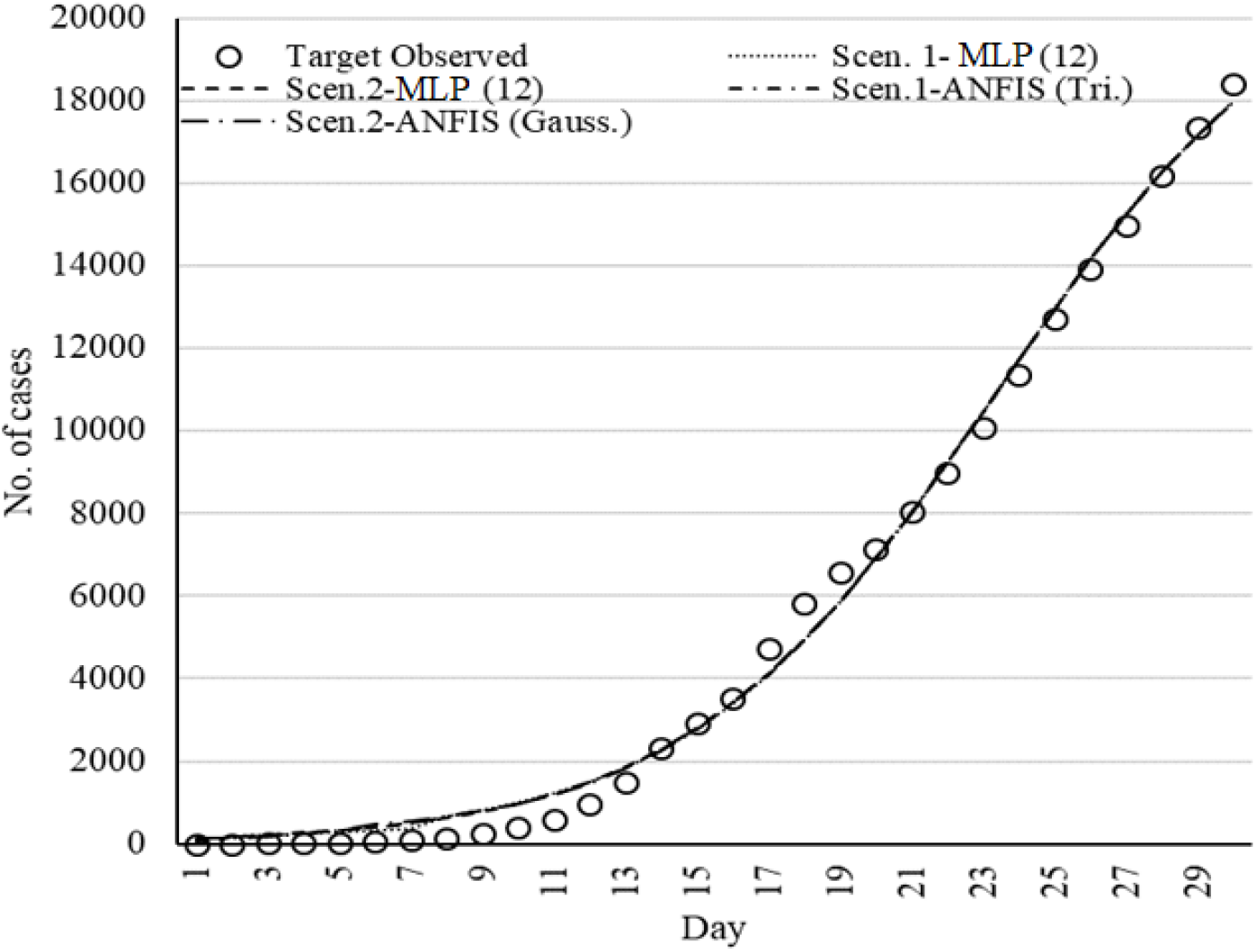
Set of models for Iran fitted by ML methods.

**Figure 15.**
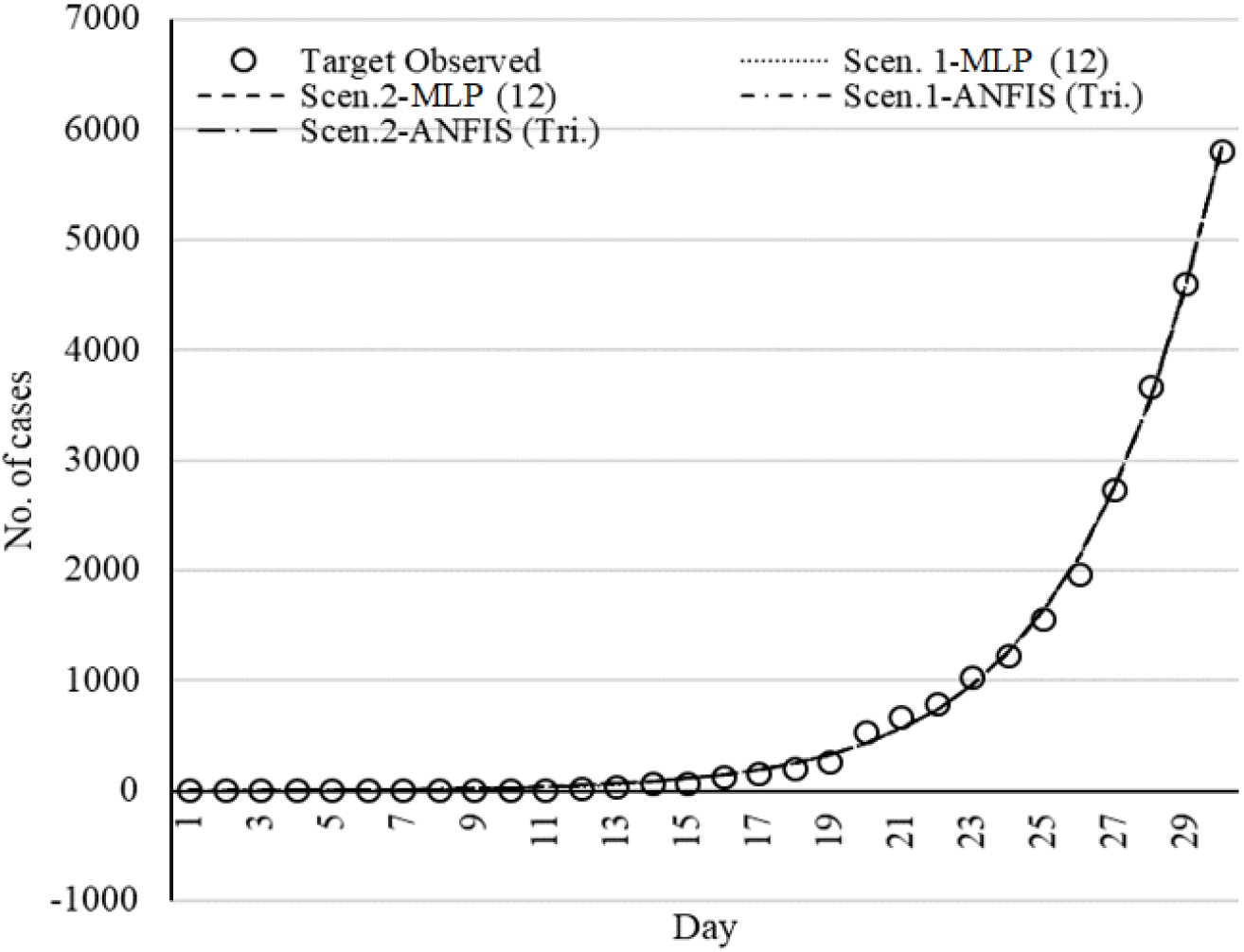
Set of models for Germany fitted by ML methods.

**Figure 16.**
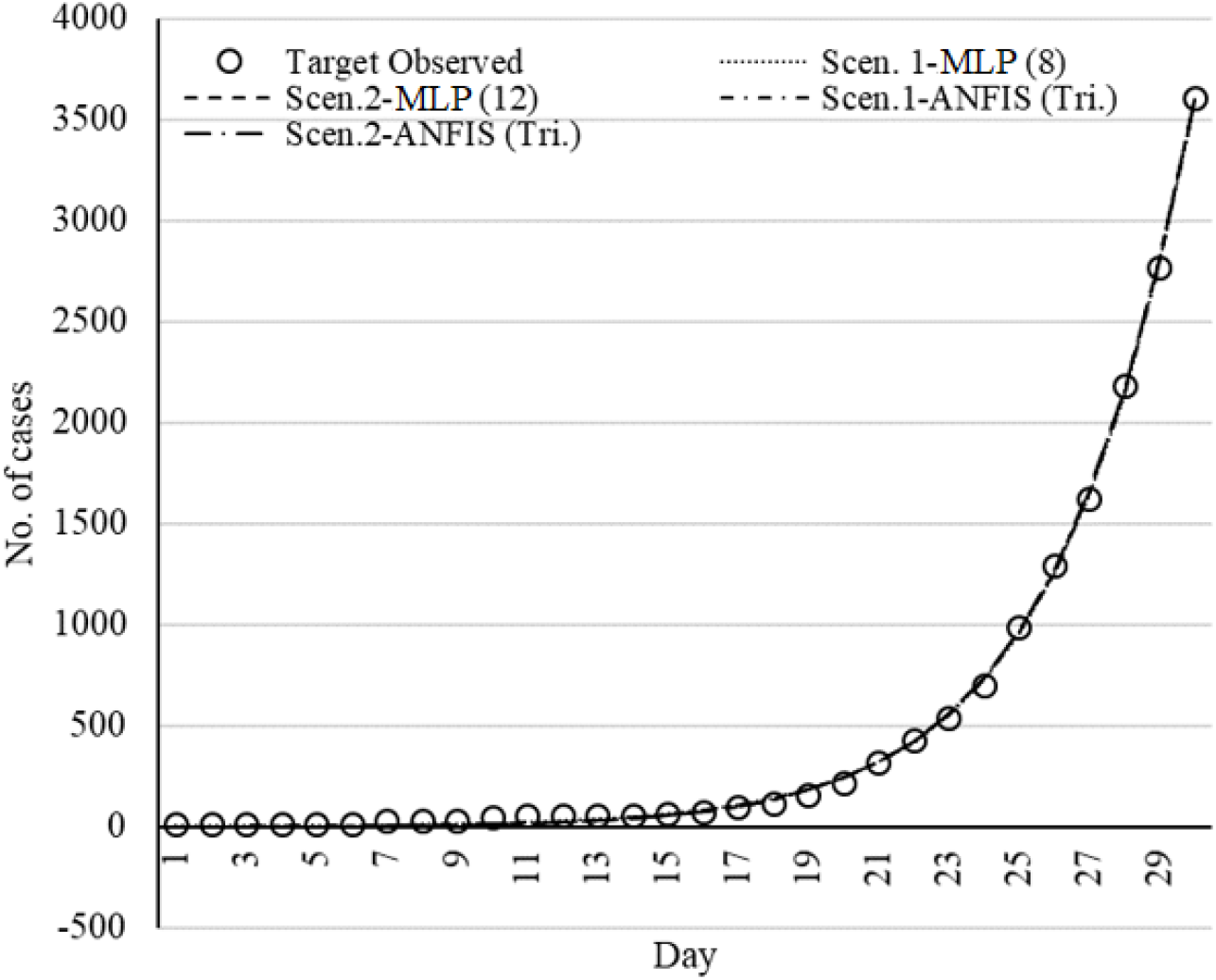
Set of models for USA fitted by ML methods.

#### Comparing the fitted models

This section presents a comparison of the accuracy and performance of the selected models for the prediction of 30 days’ outbreak. Figure 17 to 21 shows the deviation from the target values for the selected models.

**Figure 17.**
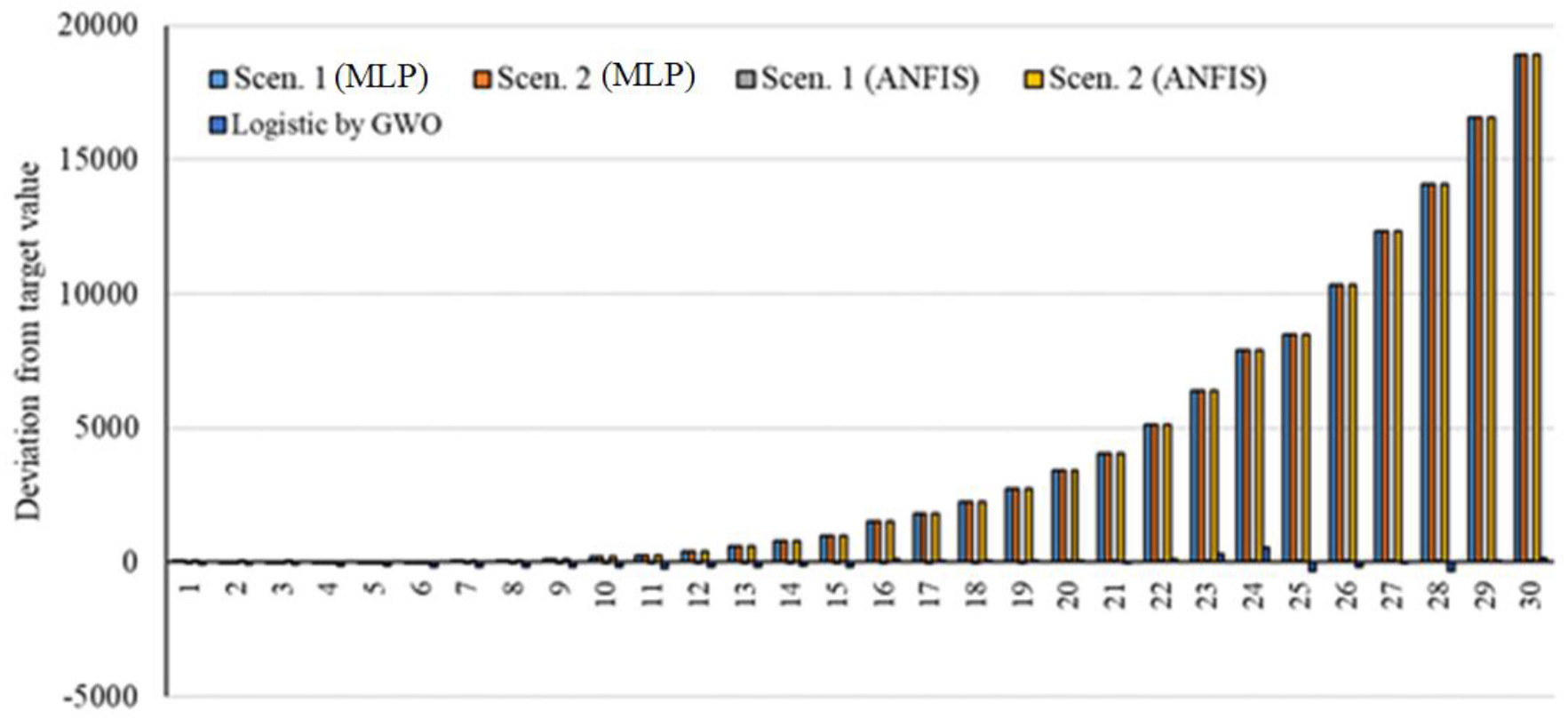
Deviation from target value for models related to Italy.

**Figure 18.**
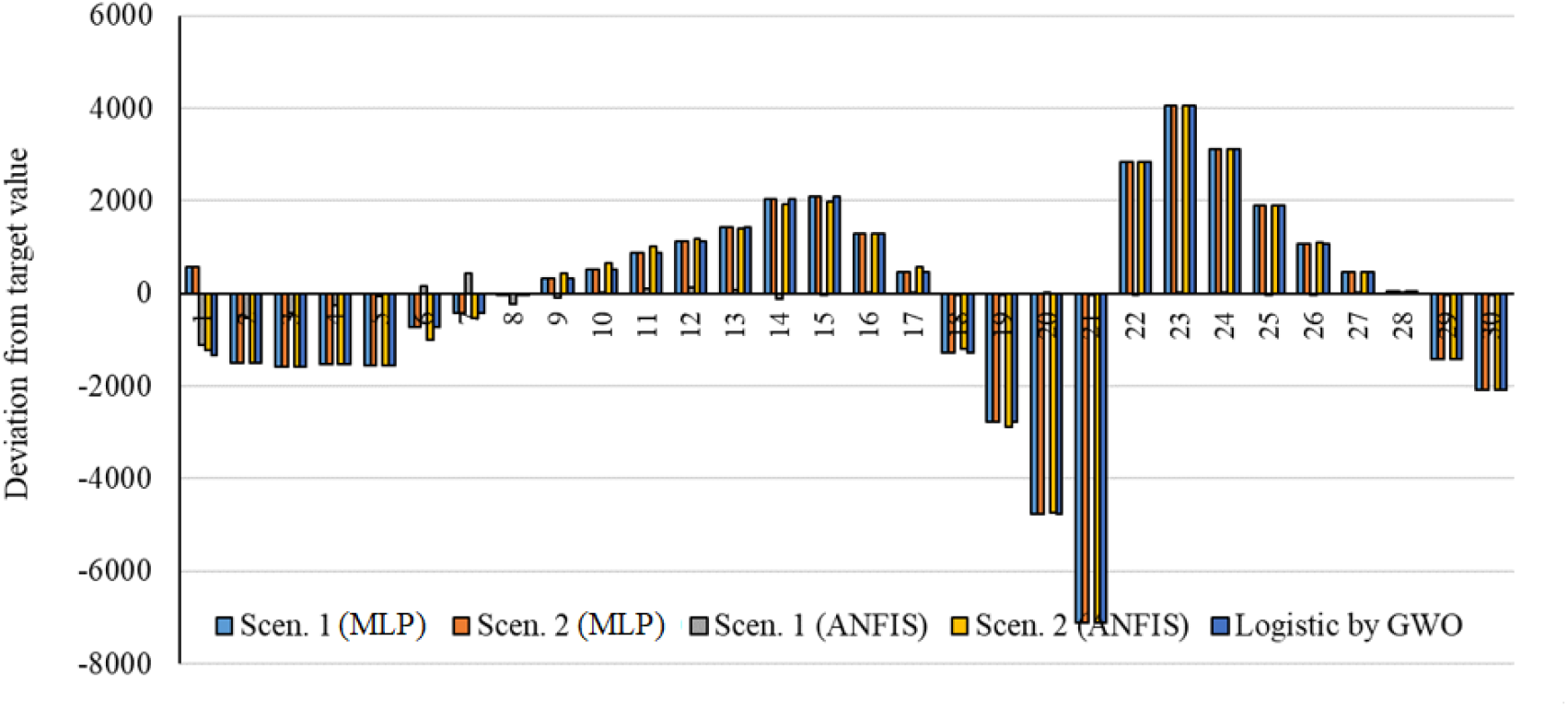
Deviation from target value for models related to China.

**Figure 19.**
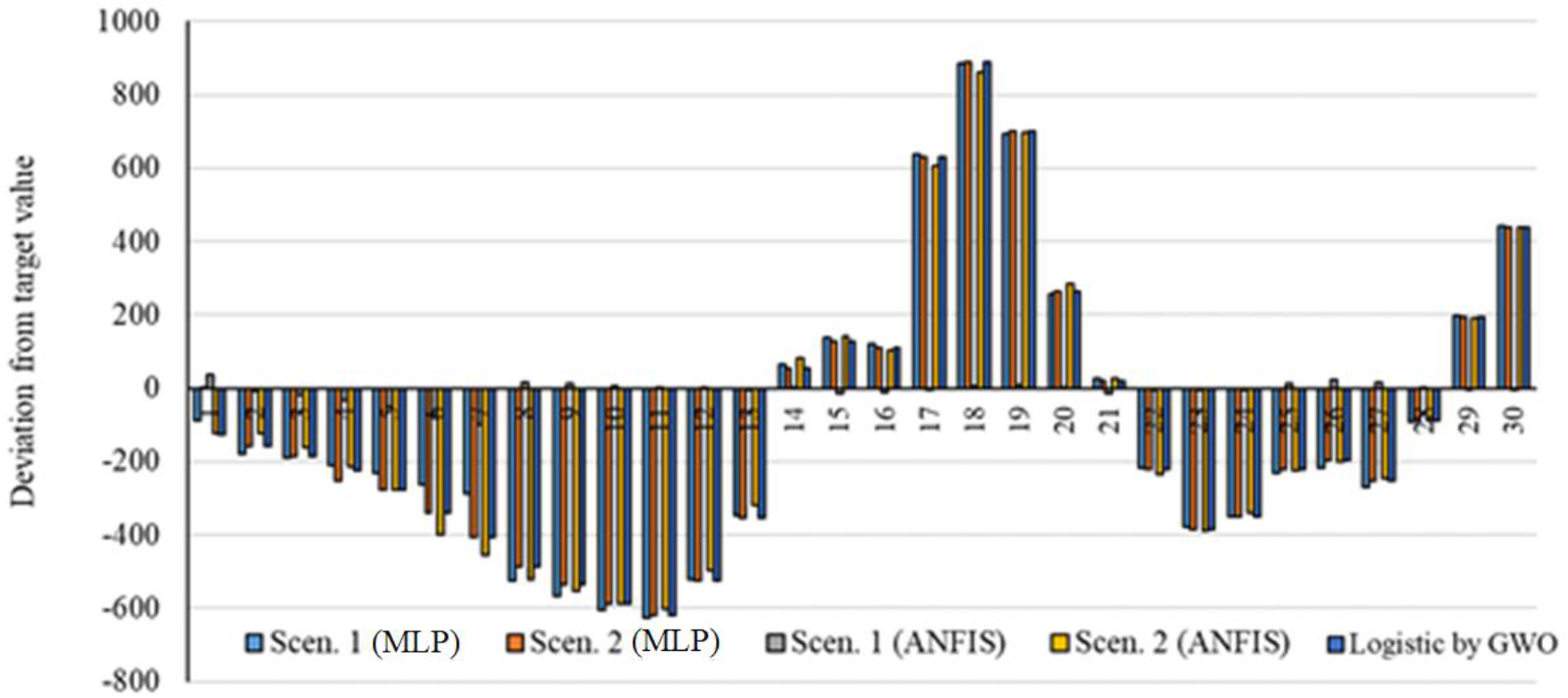
Deviation from target value for models related to Iran.

**Figure 20.**
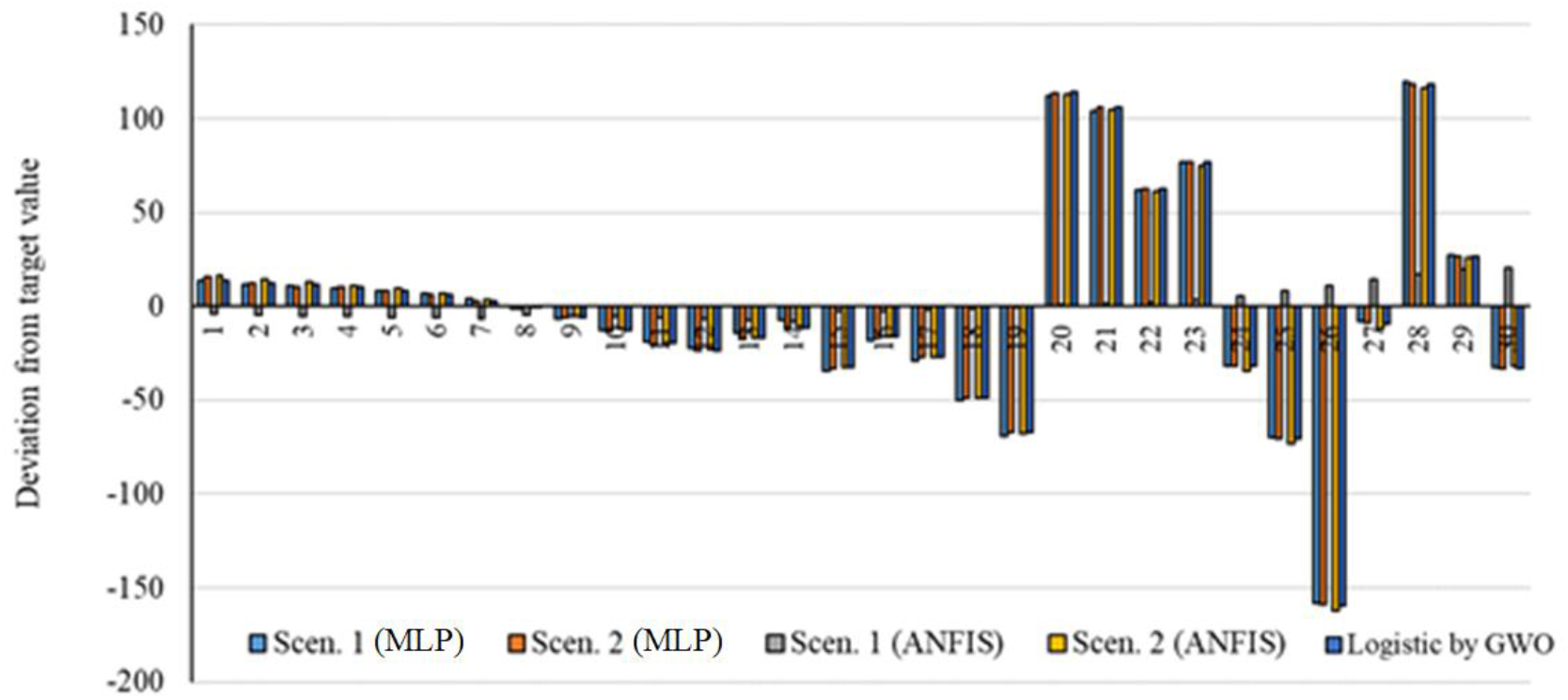
Deviation from target value for models related to Germany.

**Figure 21.**
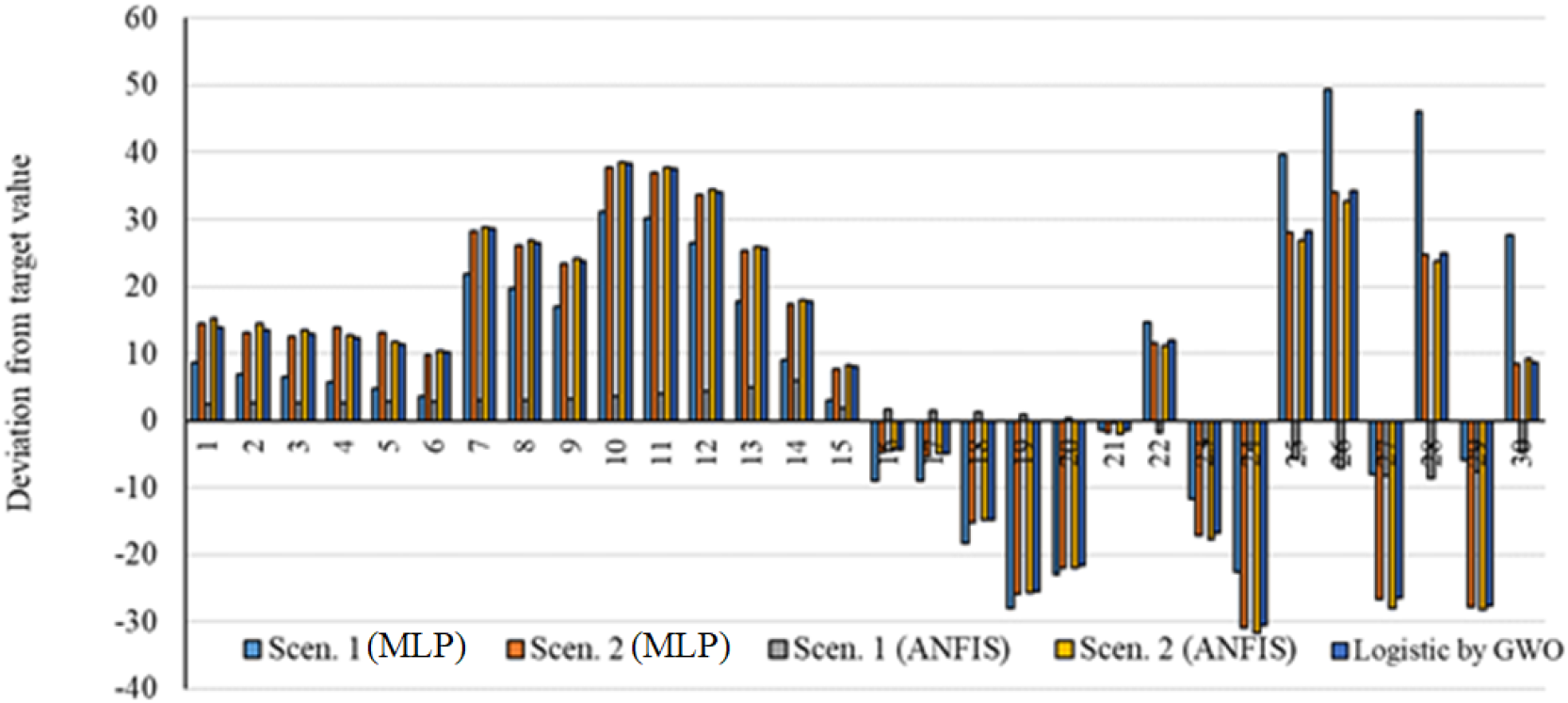
Deviation from target value for models related to USA.

As is clear from Figure 17 to 21, the smallest deviation from the target values is related to the MLP for scenario 1 followed by MLP for scenario 2. This indicates the highest performance of the MLP method for the prediction of the outbreak.

Figures 22 to 26 present the outbreak prediction for 75 days and Tables 19 to 23 present the outbreak prediction for 150 days.

**Table 19.** The outbreak prediction for Italy through 150 days

|  | Logistic<br>by GWO | Linear by<br>GWO | Logarithmic<br>by GWO | Quadratic<br>by GWO | Power by<br>GWO | MLP | ANFIS |
| --- | --- | --- | --- | --- | --- | --- | --- |
| Day<br>20th | 3794.045 | 7837.054 | -1280.93 | 5047.906 | 3225.523 | 3792.734 | 3796.738 |
| Day<br>40th | 58966.55 | 21111.37 | 273.235 | 47914.4 | 35898.08 | 58966.74 | 58964.96 |
| Day<br>60th | 70571.86 | 34385.68 | 1182.365 | 131597.7 | 146966.2 | 70571.66 | 70572.12 |
| Day<br>80th | 70729.28 | 47659.99 | 1827.402 | 256097.8 | 399523.4 | 70729.27 | 70729.15 |
| Day<br>100th | 70731.06 | 60934.31 | 2327.733 | 421414.7 | 867822 | 70731.09 | 70730.93 |
| Day<br>120th | 70731.08 | 74208.62 | 2736.532 | 627548.4 | 1635643 | 70731.14 | 70730.87 |
| Day<br>140th | 70731.08 | 87482.94 | 3082.167 | 874498.9 | 2795218 | 70731.19 | 70730.79 |
| Day<br>150th | 70731.08 | 94120.09 | 3236.862 | 1013280 | 3552851 | 70731.21 | 70730.75 |

**Table 20.** The outbreak prediction for China through 150 days

|  | Logistic<br>by GWO | Linear by<br>GWO | Logarithmic<br>by GWO | Quadratic<br>by GWO | Power by<br>GWO | MLP | ANFIS |
| --- | --- | --- | --- | --- | --- | --- | --- |
| Day 20th | 47397.6 | 47218.47 | 1341.899 | 44431.48 | 41916.55 | 47397.6 | 47360.98 |
| Day 40th | 84030.16 | 107946.8 | 9507.249 | 134729.1 | 135599.1 | 84030.17 | 84030.39 |
| Day 60th | 84996.7 | 168675.1 | 14283.67 | 265812 | 269471.3 | 84996.7 | 84996.67 |
| Day 80th | 85011.08 | 229403.4 | 17672.6 | 437680.2 | 438660.2 | 85011.08 | 85011.05 |
| Day 100th | 85011.29 | 290131.7 | 20301.26 | 650333.6 | 640132.8 | 85011.3 | 85011.22 |
| Day 120th | 85011.3 | 350860 | 22449.02 | 903772.3 | 871733.6 | 85011.34 | 85011.13 |
| Day 140th | 85011.3 | 411588.3 | 24264.94 | 1197996 | 1131815 | 85011.38 | 85011.05 |
| Day 150th | 85011.3 | 441952.5 | 25077.68 | 1360403 | 1272113 | 85011.41 | 85011.01 |

**Table 21.**
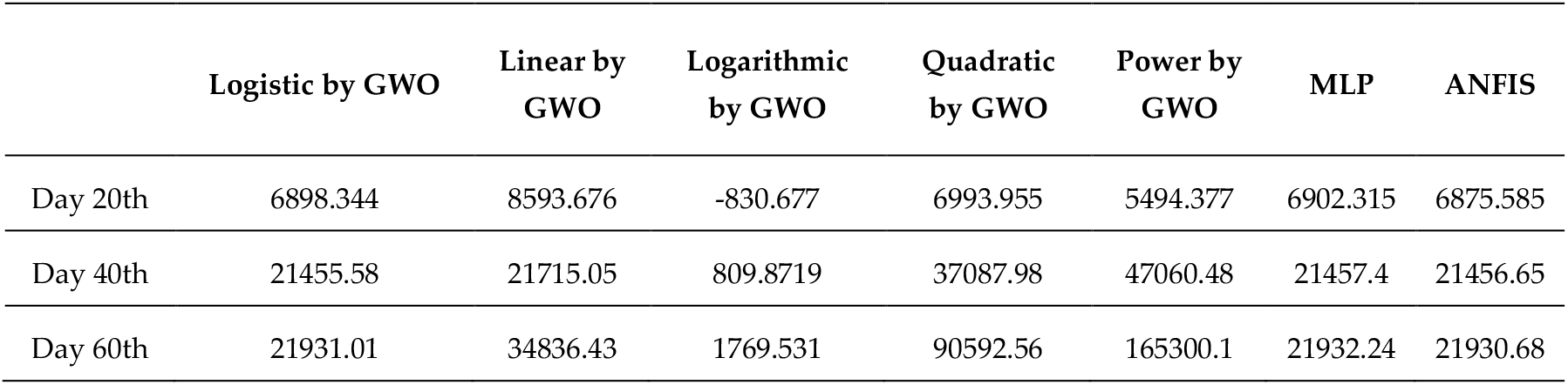
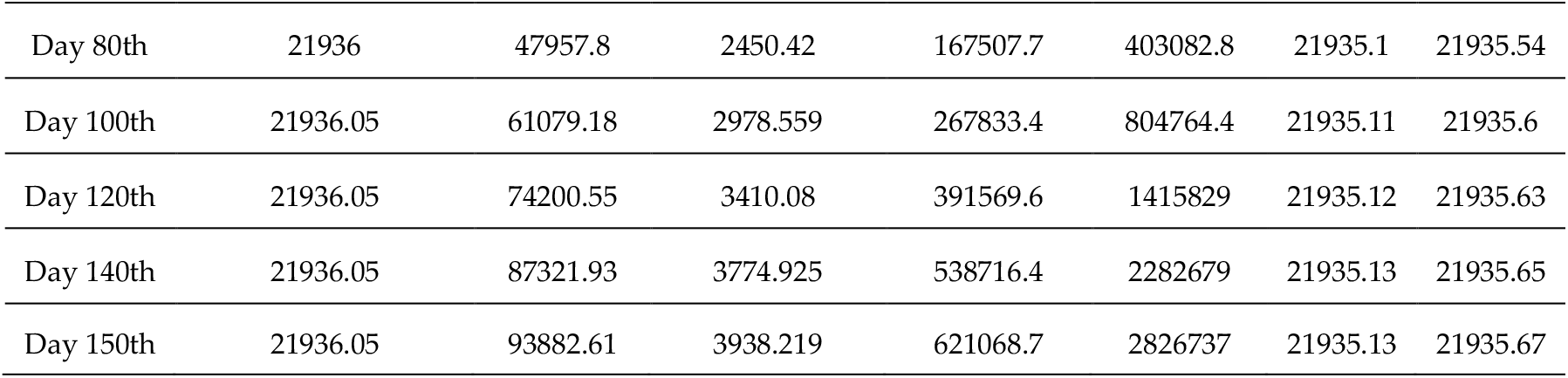
The outbreak prediction for Iran through 150 days

|  | Logistic by GWO | Linear by GWO | Logarithmic by GWO | Quadratic by GWO | Power by GWO | MLP | ANFIS |
| --- | --- | --- | --- | --- | --- | --- | --- |
| Day 20th | 6898.344 | 8593.676 | -830.677 | 6993.955 | 5494.377 | 6902.315 | 6875.585 |
| Day 40th | 21455.58 | 21715.05 | 809.8719 | 37087.98 | 47060.48 | 21457.4 | 21456.65 |
| Day 60th | 21931.01 | 34836.43 | 1769.531 | 90592.56 | 165300.1 | 21932.24 | 21930.68 |
| Day 80th | 21936 | 47957.8 | 2450.42 | 167507.7 | 403082.8 | 21935.1 | 21935.54 |
| Day 100th | 21936.05 | 61079.18 | 2978.559 | 267833.4 | 804764.4 | 21935.11 | 21935.6 |
| Day 120th | 21936.05 | 74200.55 | 3410.08 | 391569.6 | 1415829 | 21935.12 | 21935.63 |
| Day 140th | 21936.05 | 87321.93 | 3774.925 | 538716.4 | 2282679 | 21935.13 | 21935.65 |
| Day 150th | 21936.05 | 93882.61 | 3938.219 | 621068.7 | 2826737 | 21935.13 | 21935.67 |

**Table 22.**
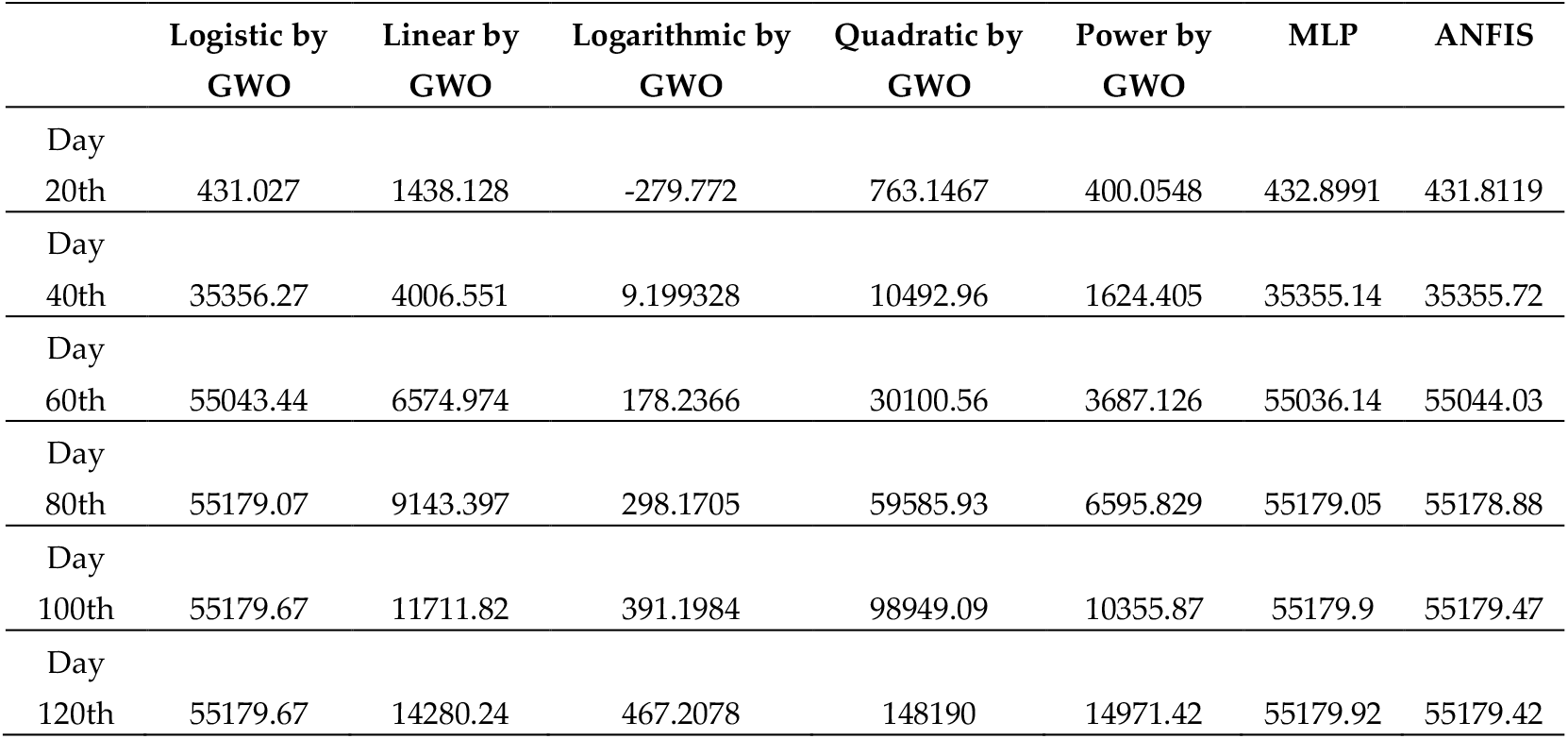
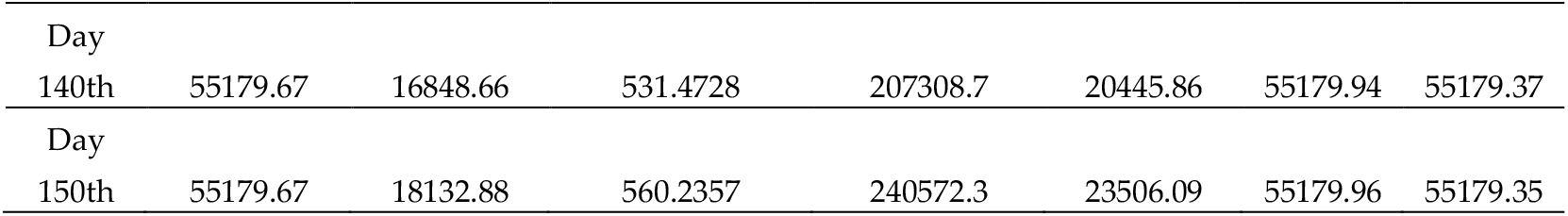
The outbreak prediction for Germany through 150 days

**Table 23.** The outbreak prediction for the USA for 150 days

|  | Logistic by<br>GWO | Linear by<br>GWO | Logarithmic by<br>GWO | Quadratic by<br>GWO | Power by<br>GWO | MLP | ANFIS |
| --- | --- | --- | --- | --- | --- | --- | --- |
| Day<br>20th | 242.6091 | 869.8855 | -156.73 | 456.0663 | 309.616 | 244.0038 | 243.6504 |
| Day<br>40th | 21951.15 | 2406.562 | 15.85698 | 6383.264 | 1031.324 | 21942.25 | 21948.25 |
| Day<br>60th | 32547.08 | 3943.238 | 116.8138 | 18366.35 | 2084.876 | 32552.6 | 32548.47 |
| Day<br>80th | 32604.34 | 5479.914 | 188.4437 | 36405.33 | 3435.319 | 32606.19 | 32604.47 |
| Day<br>100th | 32604.55 | 7016.591 | 244.0043 | 60500.21 | 5060.548 | 32606.63 | 32604.72 |
| Day<br>120th | 32604.55 | 8553.267 | 289.4005 | 90650.97 | 6944.676 | 32606.7 | 32604.76 |
| Day<br>140th | 32604.55 | 10089.94 | 327.7825 | 126857.6 | 9075.446 | 32606.78 | 32604.8 |
| Day<br>150th | 32604.55 | 10858.28 | 344.9611 | 147231.9 | 10230.16 | 32606.81 | 32604.82 |

**Figure 22.**
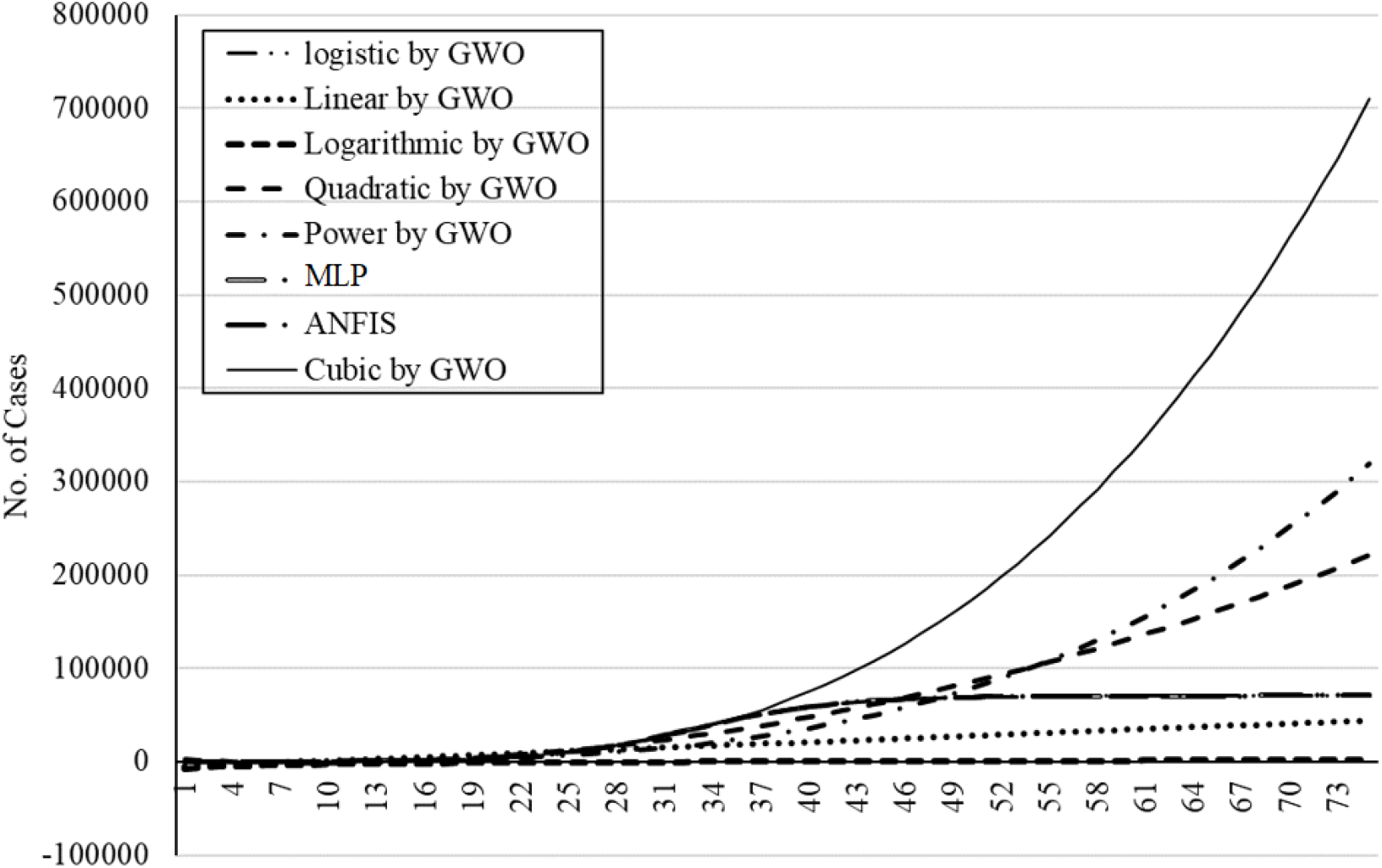
The outbreak prediction for Italy through 75 days.

**Figure 23.**
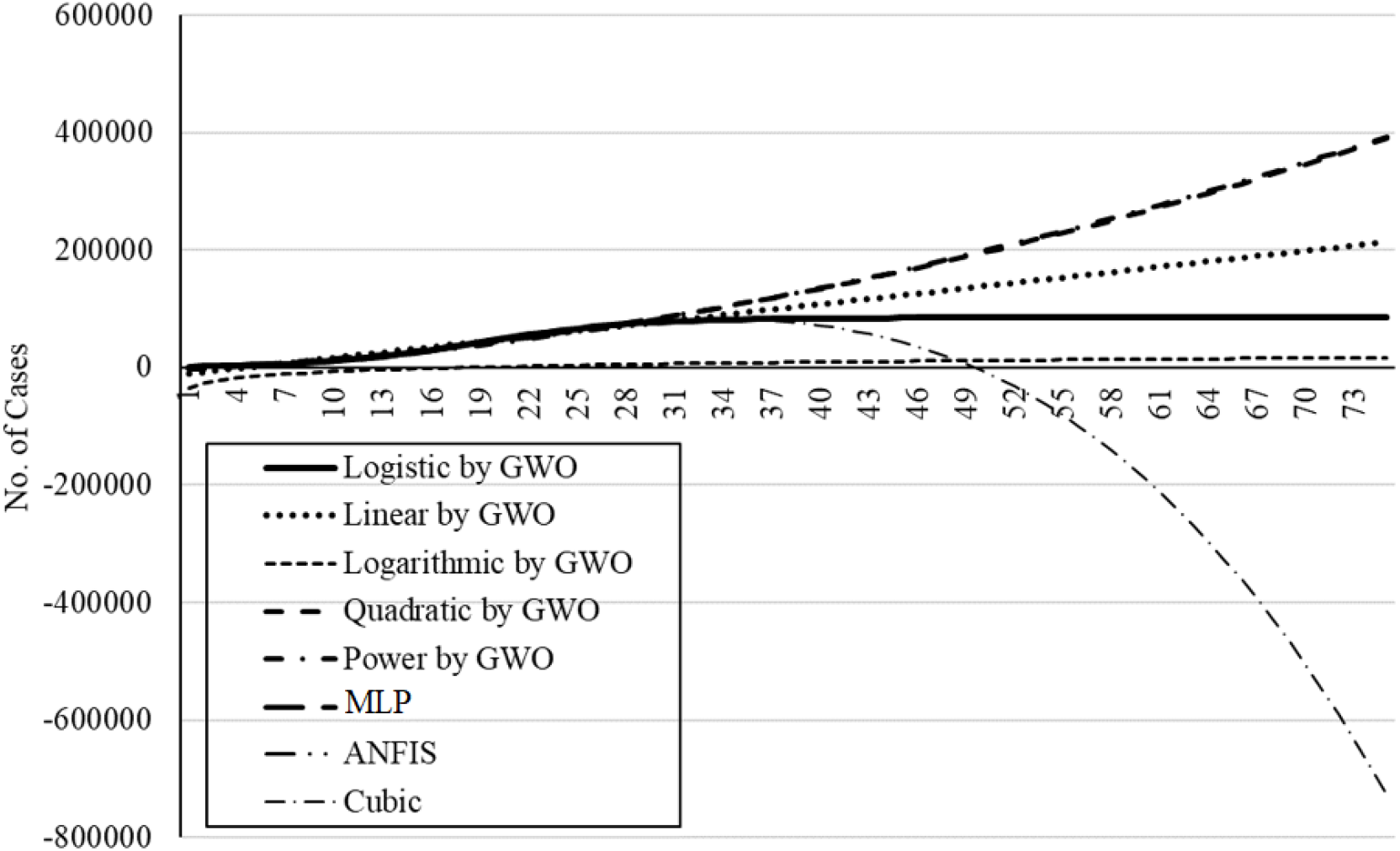
The outbreak prediction for China through 75 days.

**Figure 24.**
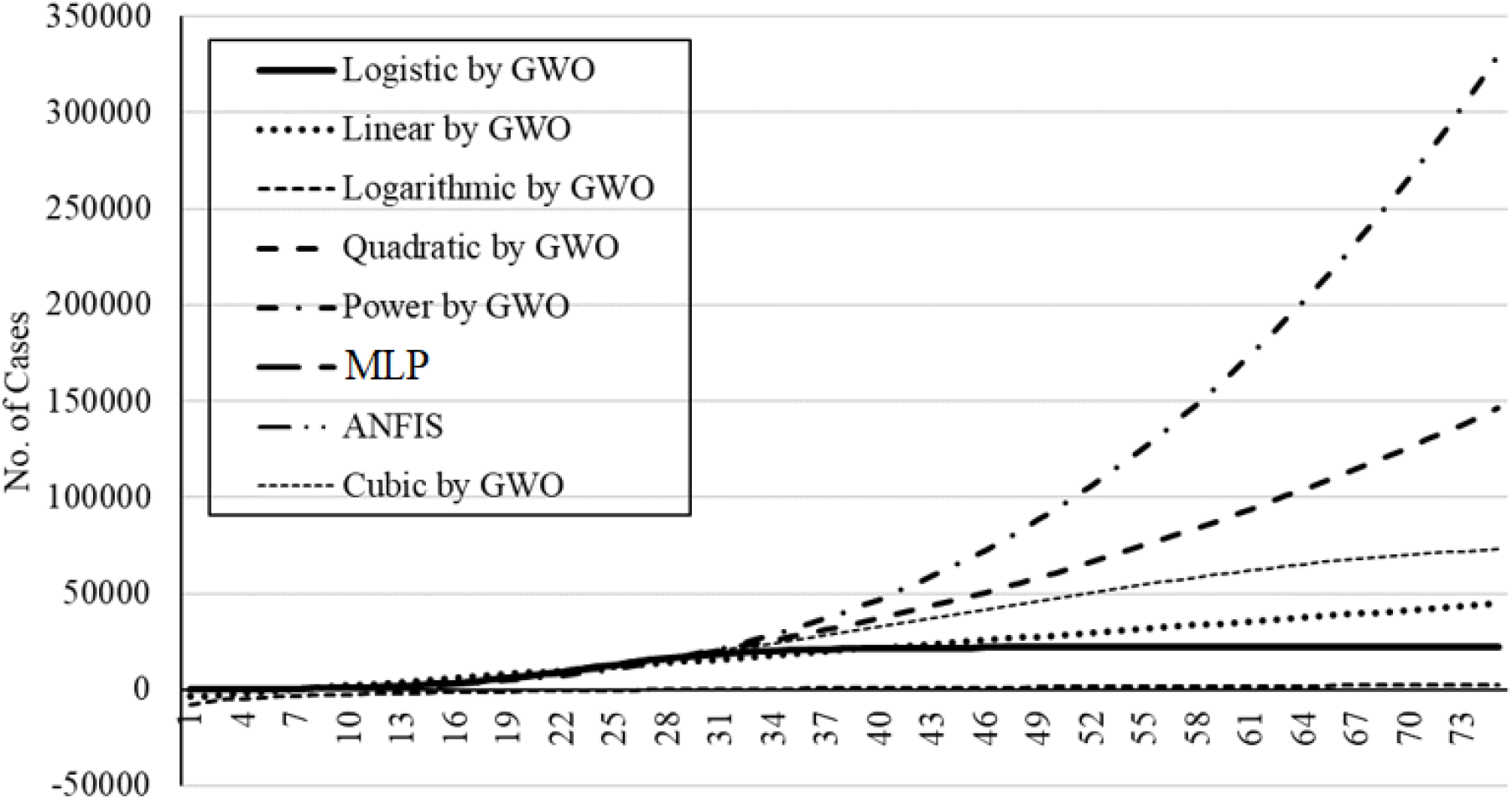
The outbreak prediction for Iran through 75 days.

**Figure 25.**
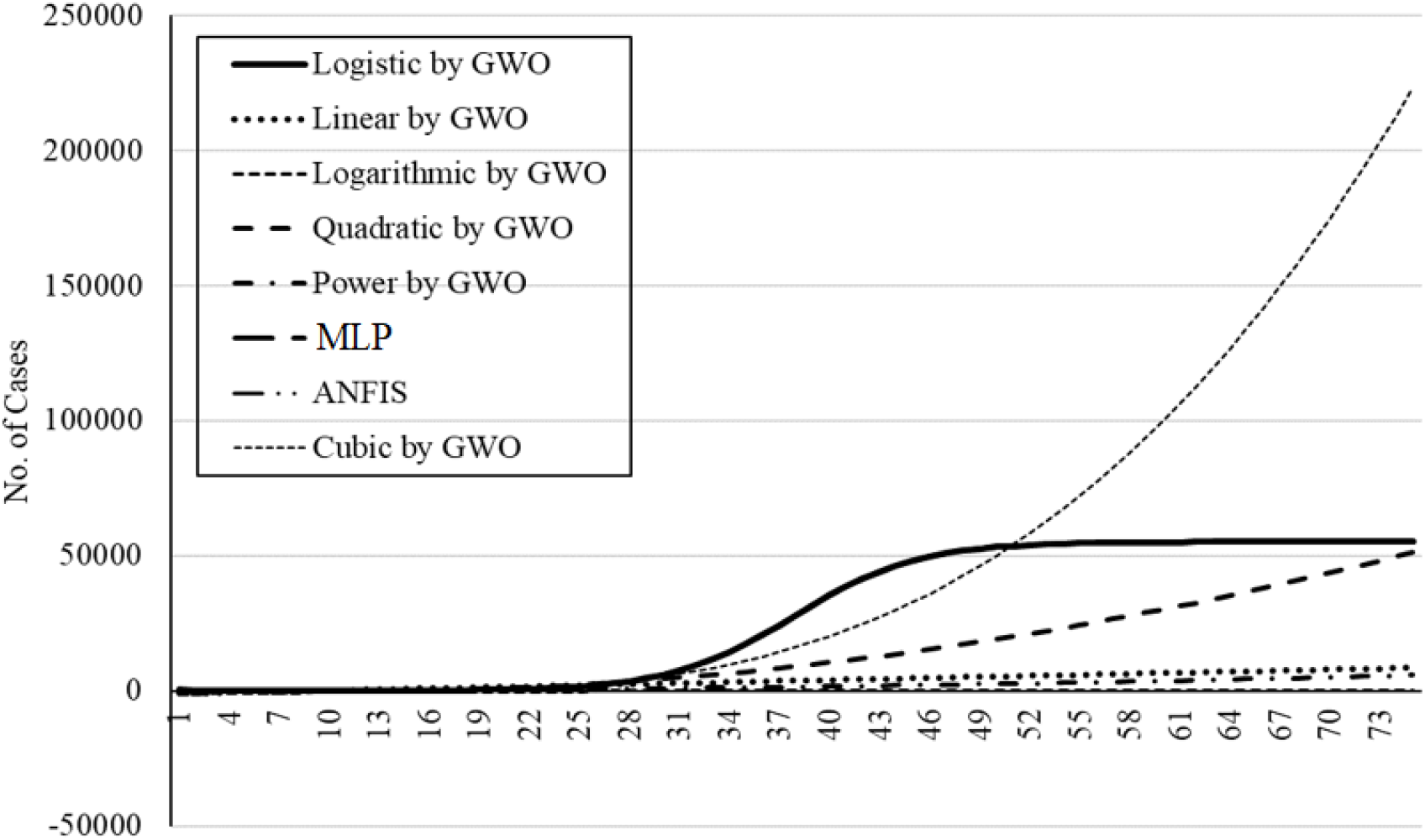
The outbreak prediction for Germany through 75 days.

**Figure 26.**
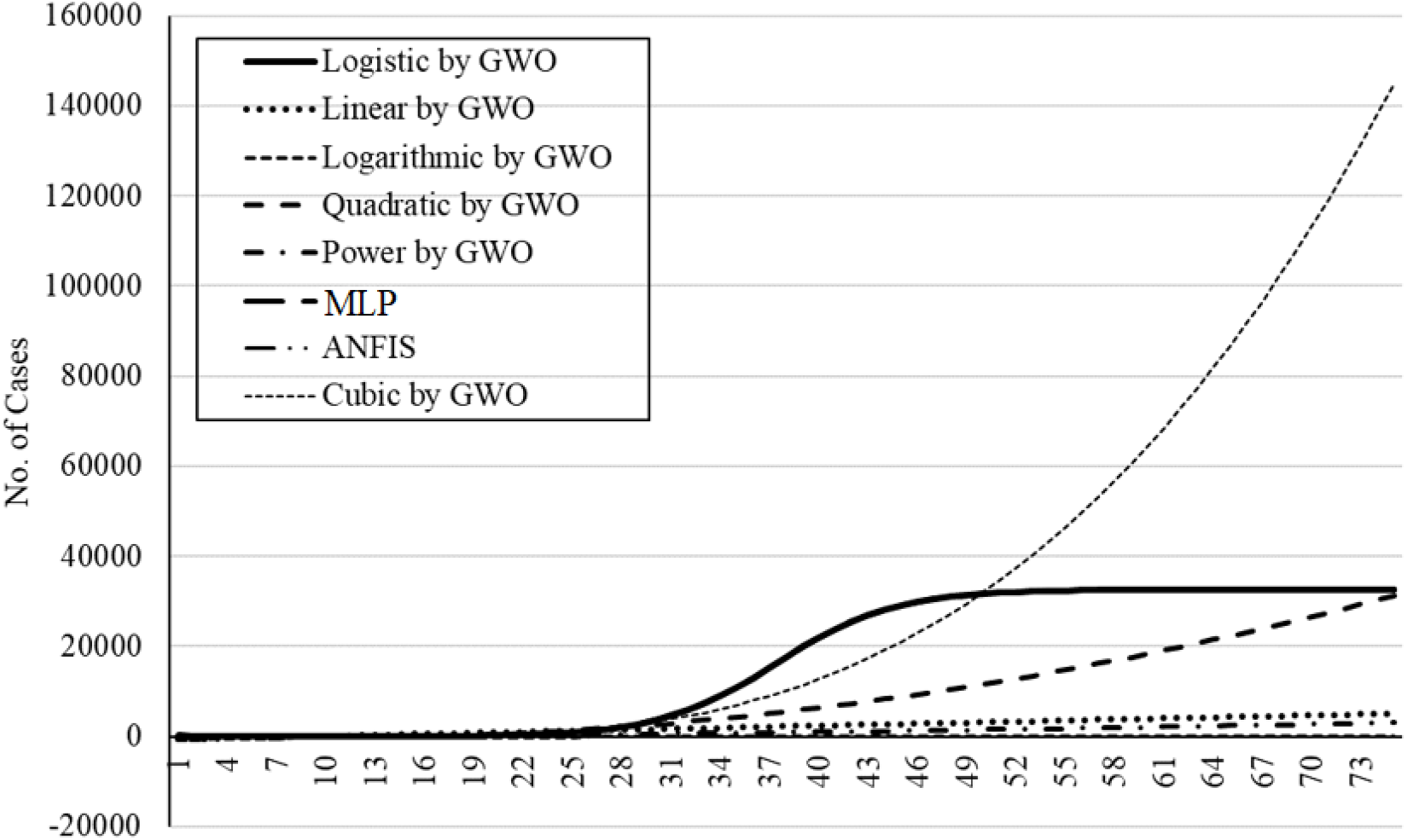
The outbreak prediction for the USA through 75 days.

## 4. Discussion

The parameters of several simple mathematical models (i.e., logistic, linear, logarithmic, quadratic, cubic, compound, power and exponential) were fitted using GA, PSO, and GWO. The logistic model outperformed other methods and showed promising results on training for 30 days. Extrapolation of the prediction beyond the original observation range of 30-days should not be expected to be realistic considering the new statistics. The fitted models generally showed low accuracy and also weak generalization ability for the five countries. Although the prediction for China was promising, the model was insufficient for extrapolation, as expected. In turn, the logistic GWO outperformed the PSO and GA and the computational cost for GWO was reported as satisfactory. Consequently, for further assessment of the ML models, the logistic model fitted with GWO was used for comparative analysis.

In the next step, for introducing the machine learning methods for time-series prediction, two scenarios were proposed. Scenario 1 considered four data samples from the progress of the infection from previous days, as reported in table 3. The sampling for data processing was done weekly for scenario 1. However, scenario 2 was devoted to daily sampling for all previous consecutive days. Providing these two scenarios expanded the scope of this study. Training and test results for the two machine learning models (MLP and ANFIS) were considered for the two scenarios. A detailed investigation was also carried out to explore the most suitable number of neurons. For the MLP, the performances of using 8, 12 and 16 neurons were analyzed throughout the study. For the ANFIS, the membership function (MF) types of Tri, Trap, and Gauss were analyzed throughout the study. The five counties of Italy, China, Iran, Germany, and USA were considered. The performance of both ML models for these countries varied amongst the two different scenarios. Given the observed results, it is not possible to select the most suitable scenario. Therefore, both daily and weekly sampling can be used in machine learning modeling. Comparison between analytical and machine learning models using the deviation from the target value (figures 17 to 21) indicated that the MLP in both scenarios delivered the most accurate results. Extrapolation for long-term prediction of up to 150 days using the ML models was tested. The actual prediction of MLP and ANFIS for the five countries was reported which showed the progression of the outbreak.

## 5. Conclusions

The global pandemic of the severe acute respiratory syndrome Coronavirus 2 (SARS-CoV-2) has become the primary national security issue of many nations. Advancement of accurate prediction models for the outbreak is essential to provide insights into the spread and consequences of this infectious disease. Due to the high level of uncertainty and lack of crucial data, standard epidemiological models have shown low accuracy for long-term prediction. This paper presents a comparative analysis of ML and soft computing models to predict the COVID-19 outbreak. The results of two ML models (MLP and ANFIS) reported a high generalization ability for long-term prediction. With respect to the results reported in this paper and due to the highly complex nature of the COVID-19 outbreak and differences from nation-to-nation, this study suggests ML as an effective tool to model the outbreak.

For the advancement of higher performance models for long-term prediction, future research should be devoted to comparative studies on various ML models for individual countries. Due to the fundamental differences between the outbreak in various countries, advancement of global models with generalization ability would not be feasible. As observed and reported in many studies, it is unlikely that an individual outbreak will be replicated elsewhere [1].

Although the most difficult prediction is to estimate the maximum number of infected patients, estimation of the *n*(deaths) / *n*(infecteds) is also essential. The mortality rate is particularly important to accurately estimate the number of patients and the required beds in intensive care units. For future research, modeling the mortality rate would be of the utmost importance for nations to plan for new facilities.

## Nomenclatures

|  |  |  |  |
| --- | --- | --- | --- |
| Multi-layered perceptron | MLP | Grey wolf optimization | GWO |
| Adaptive network-based fuzzy inference system | ANFIS | Mean square error | MSE |
| Susceptible-infected-recovered | SIR | Root mean square error | RMSE |
| Call data record | CDR | Artificial intelligence | AI |
| Classification and regression tree | CART | Artificial neural network | ANN |
| Evolutionary algorithms | EA | Triangular | Tri. |
| Genetic algorithm | GA | Gaussian | Gauss. |
| Particle swarm optimization | PSO | Trapezoidal | Trap. |
| Membership function | MF | Machine learning | ML |

## Data Availability

Data is available on request.

https://www.worldometers.info/coronavirus/country

